# A Co-infection model for HPV and Syphilis with Optimal Control and Cost-Effectiveness Analysis

**DOI:** 10.1101/2020.09.09.20191635

**Authors:** A. Omame, D. Okuonghae, U. E. Nwafor, B. U. Odionyenma

## Abstract

In this work, we develop and present a co-infection model for human papillomavirus (HPV) and syphilis with cost-effectiveness optimal control analysis. The full co-infection model is shown to undergo the phenomenon of backward bifurcation when a certain condition is satisfied. The global asymptotic stability of the disease-free equilibrium of the full model is shown not to exist, when the associated reproduction number is less than unity. The existence of endemic equilibrium of the syphilis-only sub-model is shown to exist and the global asymptotic stability of the disease-free and endemic equilibria of both the syphilis-only sub-model and HPV-only sub-model were established. The global asymptotic stability of disease-free equilibrium of the HPV-only sub-model is also proven. Numerical simulations of the optimal control model showed that the optimal control strategy which implements syphilis treatment controls for singly infected individuals is the most cost-effective of all the control strategies in reducing the burden of HPV and syphilis co-infections.

## 1 Introduction

Syphilis, caused by *Treponema Pallidum*, continues to remain a global threat to life [1]. Though treatment and condom use have proven to be effective in preventing the disease, estimates indicate that every year, about 12,000, 000 people are infected with the disease [44]. The world Health Organization (WHO) reported that about 2 million pregnant women are infected with syphilis annually [44]. Vertical transmission of syphilis can occur from a pregnant mother to her unborn child, resulting in a very negative outcomes for the pregnancy [8, 44]. The Human papillomavirus (HPV) is one of the most prevalent sexually tranmsitted infections (STIs) worldwide [3]. Globally, the Human papillomavirus is responsible for roughly 270,000 cervical cancer deaths and 97,000 other cancer deaths yearly [3]. There is no specific treatment for HPV, but the visible syptoms or genital warts due to the disease can be treated, with the goal of reducing the spread of the infection [38]. HPV, like other sexually transmitted infections, can be prevented through the use of condoms. Presently, three major anti-HPV vaccines exist: the bivalent *Cervarix* vaccine (which targets HPV-16 and –18), the quadrivalent *Gardasil 4* vaccine (which targets the oncogenic HPV types -16 and -18 and the warts causing HPV types -6 and -11) and the nonavalent *Gardasil 9* vaccine (which targets the high risk HPV types -16, -18, -31, -33, 45, -52, and -58 and the low risk HPV types -6 and -11) [43].

Epidemiological evidences have shown interactions betwen Human papillomavirus (HPV), cancer and syphilis [13, 34, 35, 45]. Souza *et al*. investigated the incidence of HPV-syphilis co-infection among patients in a hospital at Rio de Janeiro, Brazil. They discovered that many of the patients treated of HPV infections had prior syphilis cases, indicating a strong co-infection between the two infections. Zhang *et al*. [45] studied the risk factors for HPV, Human immuno-deficiency virus (HIV) and syphilis infections among men who have sexual interactions with men (MSM) in China. They discovered that HIV infection was high among individuals infected with syphilis or HPV and much higher among individuals co-infected with HPV and syphilis infections. Tseng *et al*. [35] investigated the risk factors for anal cancer in both men and women in a population-based study, and showed that prior syphilis infection was associated with persistent HPV infection and increased susceptibility to anal cancer among both genders. The findings in Daling *et al*. [13] reveal that syphilis and anal cancer co-infections were more common among unmarried men than among the married ones. In another report, da Mota et al [26] discussed the prevalence of syphilis and its risk factors among young men presented to the Brazilian Army in 2016, and showed that syphilis infection is mostly preceeded by sexually transmitted infections (STIs), especially HPV infection, gonorrhea and HIV. Similar conclusions were made by Miranda et al [25].

Mathematical modelling has extensively been applied in studying the dynamics of infectious diseases [28, 32, 40, 41]. In recent times, mathematical models have been formulated to understand the dynamics of syphilis transmission [16, 17, 27, 36]. Garnet *et al* [16] developed a mathematical model for syphilis transmission, capturing the elementary stages of the disease. They assumed infection acquired temporary immunity for infected individuals. In another paper, an SIRS (susceptible-infected-recovered-susceptible) syphilis model was fitted with real life data [17]. Iboi and Okuonghae [14] rigorously analyzed a deterministic mathematical model, incorporating early latent and late stages of syphilis infection. Saad-Roy *et al*. [36] analyzed a mathematical model for syphilis in an MSM population. More recently, Okuonghae *et al*. [27] developed a syphilis model to assess the impact of disease transmission by individuals in the early latent stage of syphilis infection on the general dynamics and transmission of the disease.

Many mathematical modelling studies have been carried out to understand the dynamics of transmission of the Human papillomavirus [4, 21, 22, 29, 37]. Malik *et al*. [22] studied the optimal control strategies for a vaccination program administering the bivalent *Cervarix*, quadrivalent *Gardasil 4*, and nonavalent *Gardasil 9* HPV vaccines to the female population. Specifically, they considered the situations where the three vaccines are used concurrently in comparison to the case where the bivalent *cervarix* and quadrivalent *Gardasil 4* vaccines were initially used and then, during the course of the vaccination program, one or two of them are interchanged with the nonavalent vaccine. Omame *et al*. [29] studied a two-sex vaccination model for HPV, to assess the imoact of condom use and treatment on the control of HPV in a population. They showed that a vaccine with 75% effectiveness rate for males and about 40% condom compliance level by females could lead to the effective control of HPV in a population.

To understand the interactions among diseases, mathematical models have been formulated and analyzed [5, 6, 24, 30, 32]. Omame *et al* [30] studied a co-infection model for HPV and tuberculosis (TB), in the presence of HPV vaccination and condom use as well as TB treatment. Their results showed that TB-only treatment strategy can significantly bring down the burden of HPV and the co-infection of the two diseases in a population. Nwankwo and Okuonghae [24] investigated the effect of syphilis treatment in a population where HIV Syphilis are co-circulating, and showed that targeted syphilis treatment could significantly curb the burden of the co-infection of the two diseases. Also, an optimal control model for two-strain tuberculosis and HIV/AIDS co-infection with cost-effectiveness analysis was studied by Agusto and Adekunle [5]. They showed that the control strategy that combines prevention of treatment failure in drug-sensitive TB infectious individuals and the treatment of individuals with drug-resistant TB was the most cost-effective in reducing the burden of the co-infection of HIV/AIDS and two strains of tuberculosis.

To the best of the authors’ knowledge, no robust optimal control mathematical model has been developed to capture the combined effect HPV vaccination and syphilis treatment on the control of HPV and syphilis co-infection, despite the availability of epidemiological evidences supporting the coinfections of both diseases. This study assesses the combined effect of these aforementioned strategies, using optimal control analysis, in order to determine ways of bringing down the bnurden of co-infection of the two diseases.

The model is based on the main assumptions stated below:

i. To avoid model complexity, the primary and secondary stages of syphilis infection are joined together and are referred to as “early stage of syphilis infection”. In a similar manner, the latent and tertiary stages of infection are referred to combined together and known as “late stage of syphilis infection”.
ii. individuals infected with syphilis infection is susceptible to infection with HPV and vice versa. [34].
iii. individuals dually infected with HPV and syphilis can transmit either HPV or syphilis but not mixed infections.

The paper is organized as follows. The model is formulated in Section 2, together with the presentation of its basic properties. The sub-models are analysed in Section 3. Qualitative and quatitative analyses of the full co-infection model with constant and time dependent controls are presented in Section 5 while Section 6 gives the concluding remarks.

## 2 Model Formulation

The total sexually active population at time *t*, denoted by *N*(*t*), is divided into twenty mutually-exclusive compartments: Susceptible individuals (*S*(*t*)), individuals in early stage of syphilis infection (*I*(*t*)), individuals in late stage of syphilis infection (*L*(*t*)), individuals treated of syphilis, (*T* (*t*)), individuals vaccinated against HPV (*V*_h_(*t*)), individuals infected with HPV (*H*(*t*)), individuals with persistent HPV infection (*P* (*t*)), individuals with anal cancer (*C*(*t*)), individuals who have recovered from anal cancer (*R*_c_(*t*)), individuals who have recovered from or cleared HPV infection (*R*_H_(*t*)), individuals dually infected with HPV and exposed to syphilis (*H*_E_(*t*)), individuals dually infected with HPV and syphilis in early stage *H*_i_(*t*), individuals dually infected with HPV and in late stage of syphilis infection, (*H*_L_(*t*)), individuals dually infected with persistent HPV and exposed to syphilis infection, (*P*_E_(*t*)), individuals dually infected with persistent HPV and in early stage of syphilis infection, (*P*_I_(*t*)), individuals dually infected with persistent HPV and in late stage of syphilis infection, (*P*_L_(*t*)), individuals dually infected with anal cancer and exposed to syphilis infection (*C*_E_(*t*)), individuals dually infected with anal cancer and in early stage of syphilis (*C*_I_(*t*)), individuals dually infected with anal cancer and in late stage of syphilis infection (*C*_L_(*t*)).

Thus

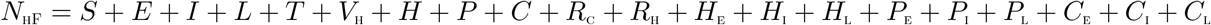

Based on the above formulations and assumptions, the Anal HPV-syphilis co-infection model is given by the following deterministic system of non-linear differential equations (the flow diagram of the model is depicted in Figure 1, the associated state variables and parameters are well described in Tables 1 and 2.

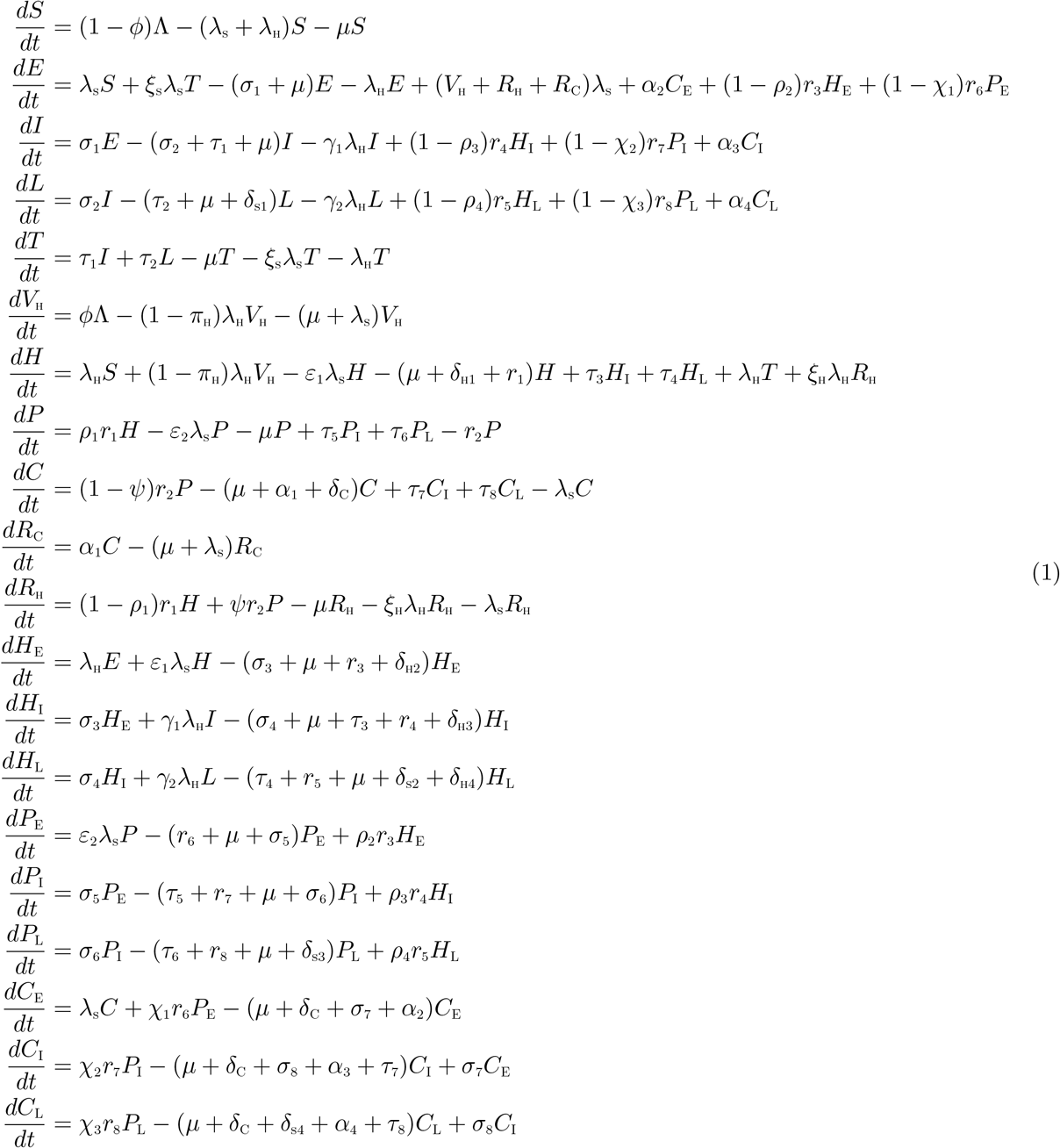

where,

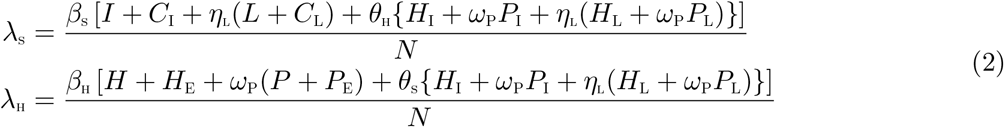

**Figure 1:**
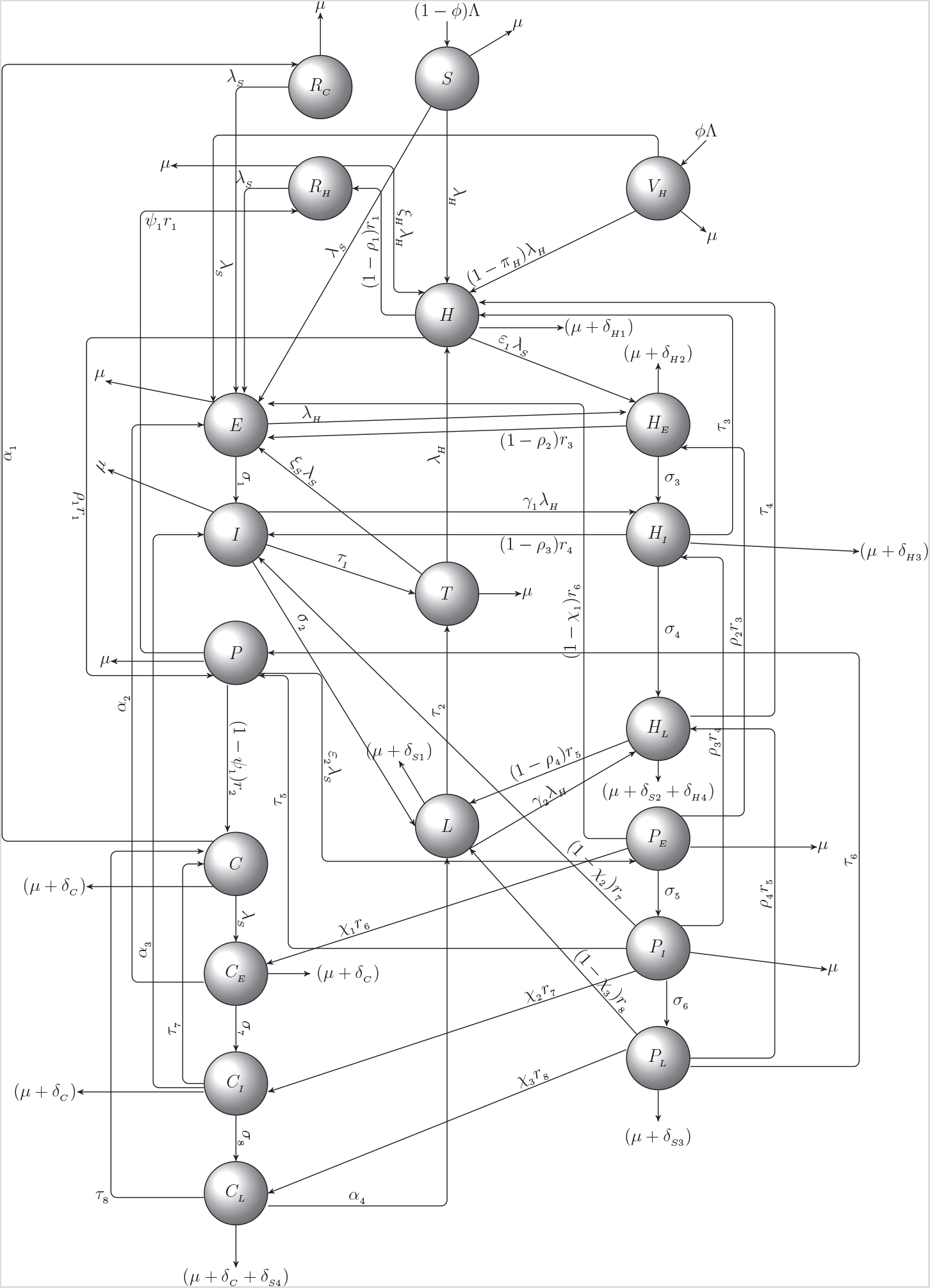
Schematic diagram of the model (1)

**Table 1:**
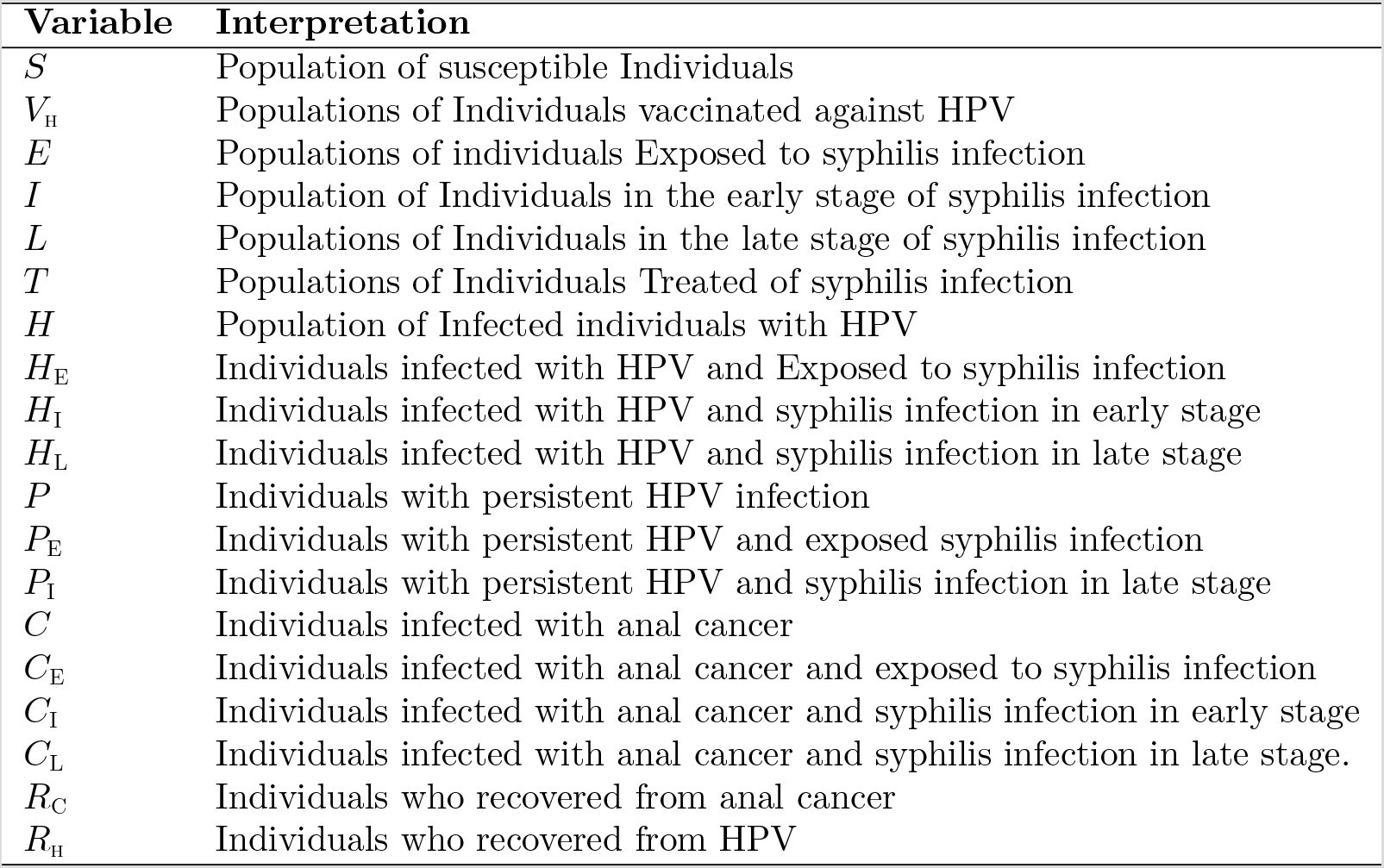
Description of variables and parameters in the model equation.

The parameter *θ*_H_(*θ*_H_ ≥ 1) is a modification parameter accounting for the increased infectiousness of dually infected individuals due to active HPV, *η*_L_(*η*_L_ ≥ 1) is a modification parameter accounting for increased infectiousness of infected individuals in the latent stage of syphilis, in comparison to those in the early stage of syphilis infection. *ω_p_*(*ω_p_ ≤* 1) is a modification parameter accounting for the reduced infectiousness of infected individuals due to persistent HPV infection while *θ*_s_(*θ*_s_ ≥ 1) is a modification parameter accounting for the increased infectiousness of dually infected individuals due to syphilis infection. In (2), *β*_s_ is the effective contact rate for the transmission of syphilis infection, while, *β*_H_ is the effective contact rate for the transmission of HPV infection.

### 2.1 Basic properties of the co-infection model (1)

The basic dynamical properties of the model (1) will now be explored. Particularly, we establish the following positivity and invariance results.

#### 2.1.1 Positivity and boundedness of solutions

For the model (1) to be epidemiologically meaningful, it is important to prove that all its state variables are non-negative for all time. Following the same approach in Omame *et al* [29], we establish the following results.

**Theorem 2.1** *Let the initial data be*

*S*(0) > 0, *E*(0) ≥ 0, *I*(0) ≥ 0, *L*(0) ≥ 0, *T* (0) ≥ 0, *V*_H_(0) ≥ 0, *H*(0) ≥ 0, *P* (0) ≥ 0, *C*(0) ≥ 0, *R*_C_(0) ≥ 0, *R*_H_(0) ≥ 0, *H*_E_(0) ≥ 0, *H*_I_(0) ≥ 0, *H*_L_(0) ≥ 0, *P*_E_(0) ≥ 0, *P*_I_(0) ≥ 0, *P*_L_(0) > 0, *C*_E_(0) ≥ 0, *C*_I_(0) ≥ 0, *C*_L_(0) ≥ 0.

*Then the solutions*

(*S, E, I, L, T, V*_H_, *H, P, C, R*_C_, *R*_H_, *H*_E_, *H*_I_, *H*_L_, *P*_E_, *P*_I_, *P*_L_, *C*_E_, *C*_I_, *C*_L_) *of the model* (1) *are non-negative for all time t >* 0.

**Lemma 2.1** *The region* 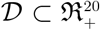 *is positively-invariant for the co-infection model* (1) *with initial conditions in* 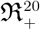.

Therefore, 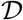 is positively invariant. Hence, no solution path can leave through any boundary of 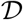 and it is sufficient to consider the dynamics of the model (1) in 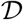. Inside this region, the model is considered to be mathematically and epidemiologically well posed.

## 3 Analysis of the sub-models

It is instructive to analyze the sub-models of the co-infection model (1), before analyzing the full model.

### 3.1 Syphilis-Only sub-model

The syphilis-only sub-model is (obtained by setting *V*_H_ = *H* = *P* = *C* = *R*_c_ = *R*_H_ = *H*_E_ = *H*_I_ = *H*_L_ = *P*_E_ = *P*_I_ = *P*_L_ = *C*_E_ = *C*_I_ = *C*_L_ = 0 in the model (1)) given by:

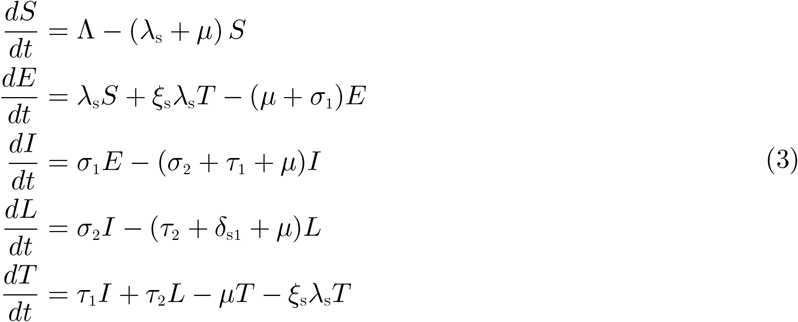

where now,

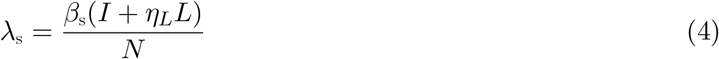

with

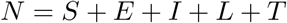

#### 3.1.1 Basic reproduction number of the syphilis only sub-model

The syphilis-only sub-model (3) has a DFE, obtained by setting the right-hand sides of the equations in the model (3) to zero, given by

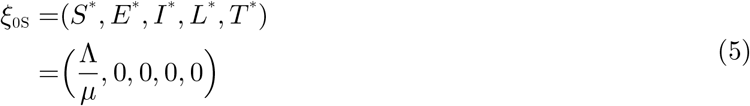

The basic reproduction number, using the next generation operator method [42], is given by

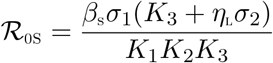

with,

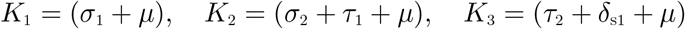

#### 3.1.2 Local asymptotic stability of disease-free equilibrium (DFE) of the syphilis-only submodel

**Lemma 3.1** *The DFE, ξ*_0_*S, of the syphilis-only sub-model* (3) *is locally asymptotically stable (LAS) if* 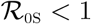 and unstable if 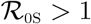.

**Proof:**

The local stability of the syphilis-only sub-model is analysed by the Jacobian matrix of the system (3) at *ξ*_0S_, given by:

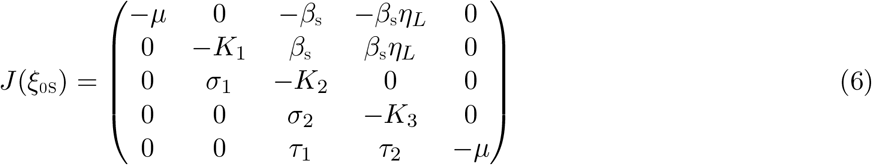

where,

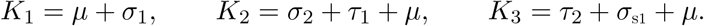

The eigenvalues are *−µ, −µ* and the solution of the characteristic polynomial:

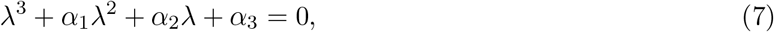

where

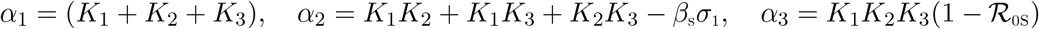

Applying the Routh-Hurwitz criterion, the cubic equation (7) will have roots with negative real parts if and only if *α*_1_ > 0, *α*_3_ > 0 and *α*_1_*α*_2_ > *α*_3_. Clearly, *α*_1_ > 0 and *α*_3_ > 0 (if 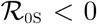). As a result, the disease-free equilibrium, *ξ*_0S_ is locally asymptotically stable if 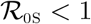.

#### 3.1.3 Global asymptotic stability(GAS) of the disease-free equilibrium(DFE) *ξ*_0*S*_ of the syphilis-only sub-model

We use the method presented in Castillo-Chavez *et al* [11] to investigate the global asymptotic stability of the disease free equilibrium of the syphilis-only sub-model. In this section, we list two conditions that if met, also guarantee the global asymptotic stability of the disease-free state. First, system (3) must be written in the form:

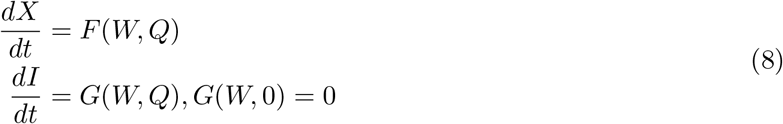

where *W* ∈ *R^m^* denotes (its components) the number of uninfected individuals and *Q ∈ R^n^* denotes (its components) the number of infected individuals including latent, infectious, etc. *U*_0_ = (*W^∗^*, 0) denotes the disease-free equilibrium of this system. The conditions (*H*1) and (*H*2) below must be met to guarantee local asymptotic stability:

(*H*1): For 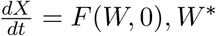 is globally asymptotically stable (GAS),
(*H*2): 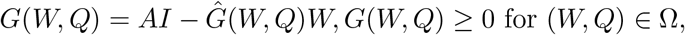

where *A* = *D*_1_*G*(*W^∗^*, 0) is an M-matrix (the off-diagonal elements of *A* are nonnegative) and Ω is the region where the model makes biological sense. If System (3) satisfies the above two conditions then the following theorem holds:

**Theorem 3.1** *The fixed point U*_0*S*_ = (*W^∗^*, 0) *is a globally asymptotic stable (GAS) equilibrium of* (3) *provided that R*_0_ < 1 *(LAS) and that assumptions* (*H*1) *and* (*H*2) *are satisfied*

**Proof**

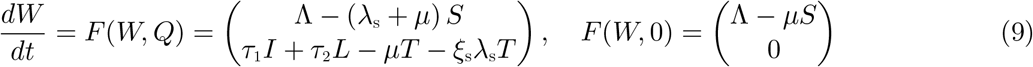

where W denotes the number of non-infectious individuals and Q denotes the number of infected individuals

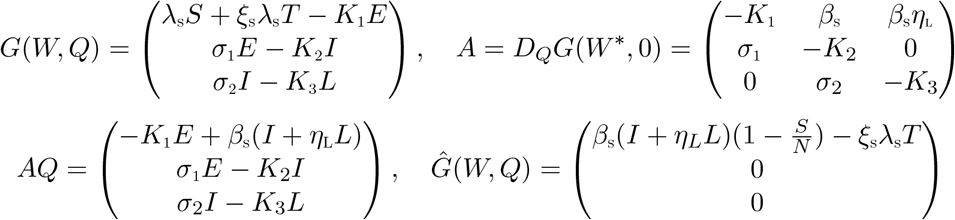

It is clear from the above, that, 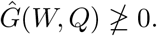. Hence the DFE, *U*_0*S*_, may not be globally asymptotically stable, suggesting the possibility of a backward bifurcation. This supports the backward bifurcation analysis in Section (3.1.5)

#### 3.1.4 Endemic equilibrium point (EEP) of the syphilis-only sub-model

In this section, we obtain the endemic equilibrium point (EEP) of the syphilis-only sub-model (3). The endemic quilibrium point (EEP) of the syphilis-only sub-model (3) is given by

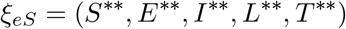

where,

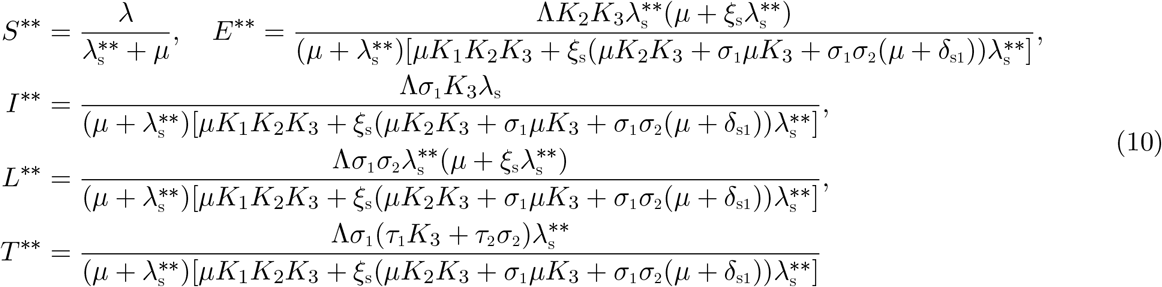

substituting the right-hand sides of (10) into the force of infection (4) at steady states, we obtain the polynomial equation

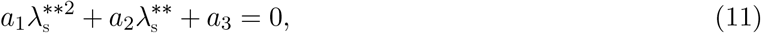

where,

*a*_1_ = (*K*_2_*K*_3_ + *σ*_1_*K*_3_ + *σ*_1_*σ*_2_)*ξ*_s_,
*a*_2_ = (*µK*_2_*K*_3_ + *σ*_1_*σ*_2_(*µ* + *δ*_s1_) + *σ*_1_*µK*_3_)*ξ*_s_ + *µK*_2_*K*_3_ + *µσ*_1_*K*_3_ + *µσ*_1_*σ*_2_ + *σ*_1_(*τ*_1_*K*_3_ + *τ*_2_*σ*_2_) *− β*_s_*σ*_1_*ξ*_s_(*K*_3_ + *η*_L_*σ*_2_)
*a*_3_ = *µK*_1_*K*_2_*K*_3_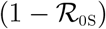

It is worthy of note, from the equation (11), that *a*_1_ > 0, while *a*_3_ is greater than zero (less than zero) if 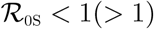. Hence, the following result can be established:

**Theorem 3.2** *The model* (3) *has*

i. *a unique endemic equilibrium if* 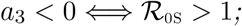
ii. *a unique endemic equilibrium if a*_2_ < 0 *and a*_1_ = 0 *or* 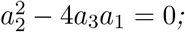
iii. *two endemic equilibria if a*_1_ > 0, *a*_2_ < 0 *and* 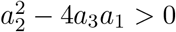 *and* 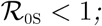
iv. *no endemic equilibrium otherwise*.

It is also interesting to note that, setting the re-infection term *ξ*_s_ = 0, reduces the quadratic (11) to 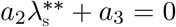, resulting in no sign changes in the polynomial equation (11), as *a*_2_ > 0 and *a*_3_ > 0 (for 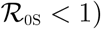. Hence no existence of an endemic equilibrium for 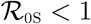, ruling out the existence of backward bifurcation in the syphilis only sub-model (3) in the absence of re-infection of treated individuals. This is consitent with the proof in Theorem 3.1

#### 3.1.5 Bifurcation analysis of the syphilis-only sub-model

Using the same approach as in Castillo-Chavez and Song [10], we shall investigate the possibility of a backward bifurcation for the syphilis-only sub-model. The linearized system evaluate at the DFE is given as

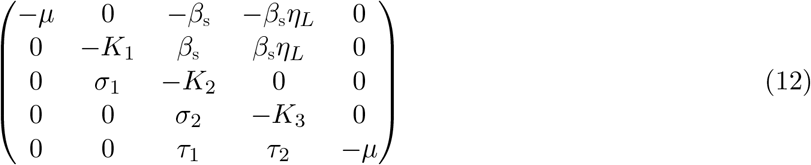

where

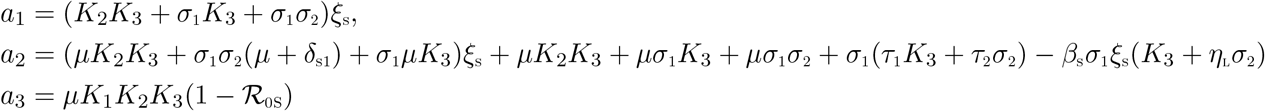

The components of the right eigenvector are given as:

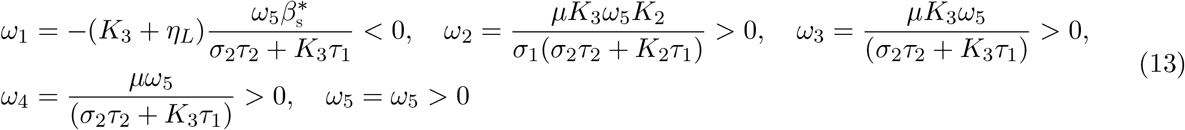

The components of the left eigenvector are equally given by:

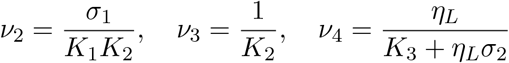

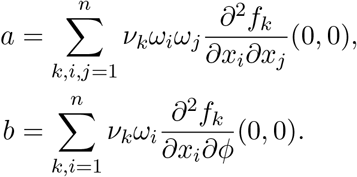

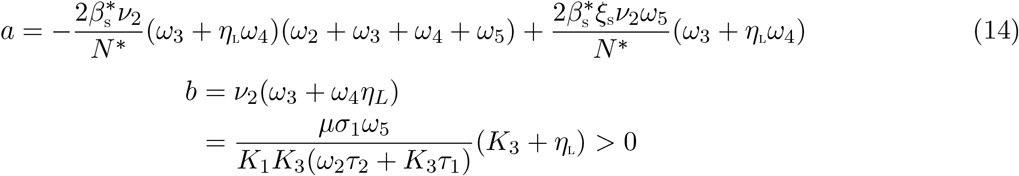

It can be observed from (14), that setting the re-infection parameter, *ξ*_S_ = 0, results in *a* < 0. Thus, re-infection induced the phenomenon of backward bifurcation in the Syphilis-only sub-model (3). This is consistent with the results obtained in the analyses in sections 3.1 and 3.1.4. An associated backward bifurcation diagram is depicted in Figure 2).

**Figure 2:**
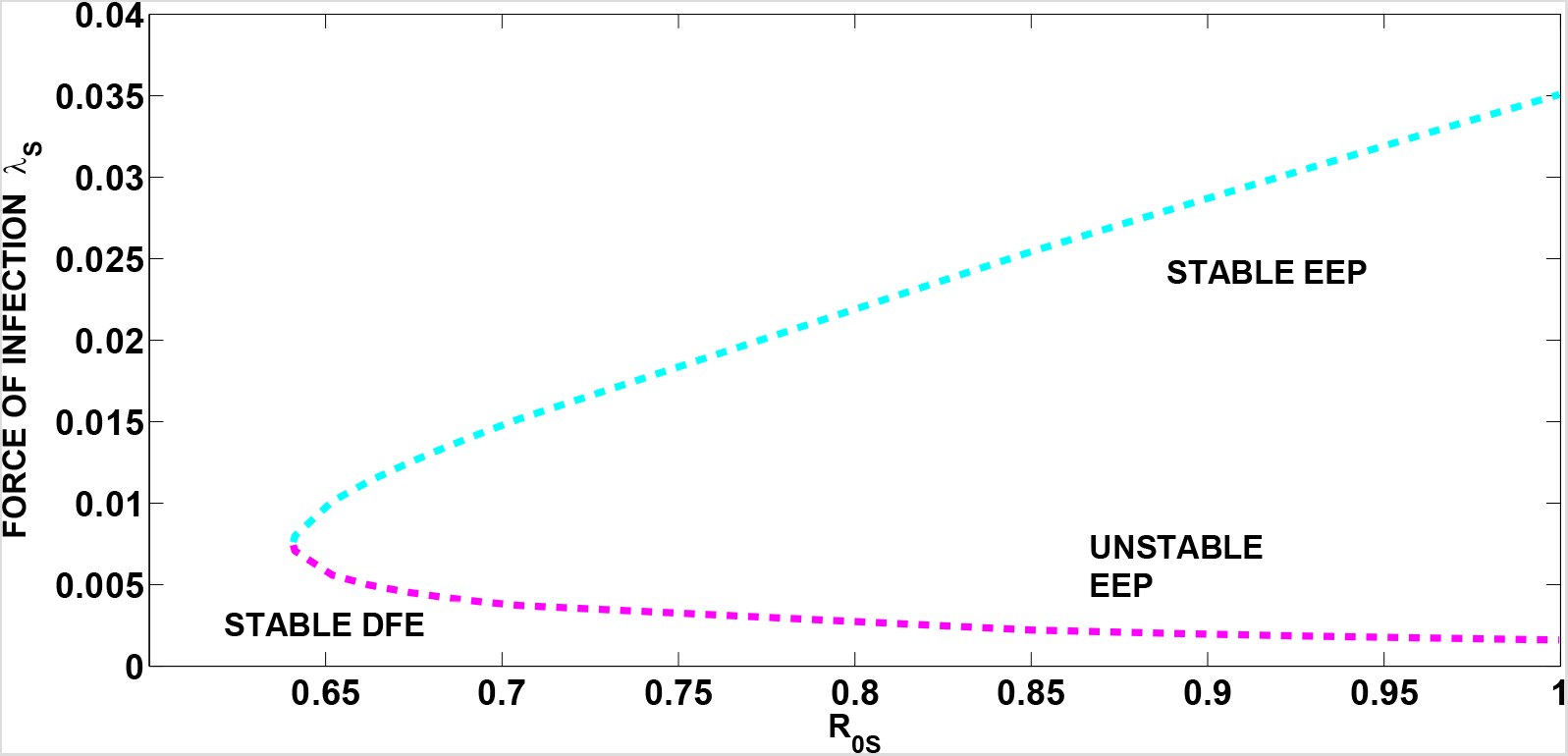
Bifurcation diagram for the syphilis-only sub-model (3). Parameter values used are: *β*_s_ = 3.0129, *ξ*_s_ = 6.0. All other parameters as in Table 2

#### 3.1.6 GAS of EEP of the Syphilis-only sub-model (3): special case(*ξ*_s_ = *δ*_s_ = 0)

Now that the cause of the backward bifurcation in the syphilis-only sub-model (3) is removed, that is, *ξ*_s_ = 0, we seek to prove the global asymptotic stability of the unique endemic equilibrium of the sub-model, for a special case when disease induced death rates are neglibible, that is, *δ*_s_ = 0.

The Syphilis-only sub-model (3) has an endemic equilibrium given by

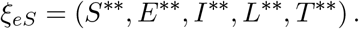

It should be noted that setting *δ*_s_ = 0 in (3) gives us 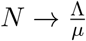 as *t* → ∝. Let 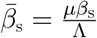 so that

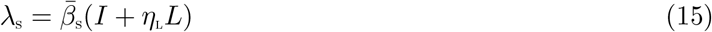

**Theorem 3.3** *Consider the sub-model* (3) *with ξ*_s_ = 0*. The sub-model is GAS in* 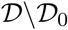 *whenever* 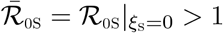, *where*

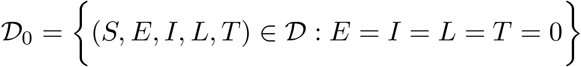

*Proof*. Consider the syphilis-only sub-model (3) with (15) and *ξ*_s_ = 0 and 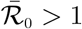, so that the associated unique endemic equilibrium exists. Also, consider the Lyapunov functional similar to the Goh-Volterra type considered by Ghosh et al [15]:

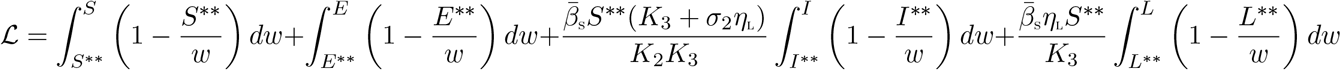

The time derivative, using Leibniz rule for integration as illustrated in Adams [2], is given by

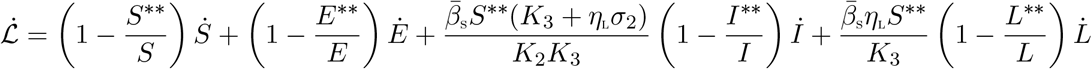

Substituting the expressions for the derivatives, 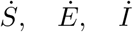 and 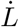 from (3), into the Lyapunov derivative, 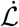, and carrying out certain algebraic manipulations, we have that

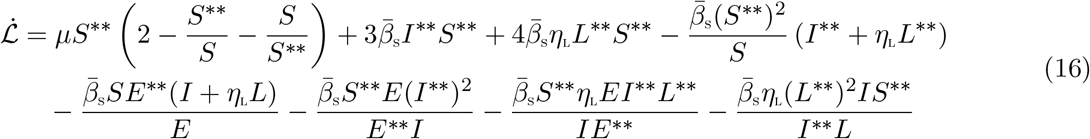

(16) can be further simplified into

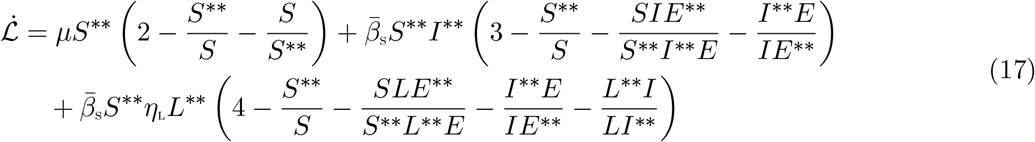

Thus, 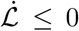 for 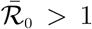. Hence, 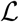 is a Lyapunov function in 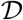 and it follows from the La Salle’s Invariance principle [19], that every solution to the equations of the syphilis-only sub-model (3) with (15) and initial conditions in 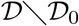 approaches the associated unique endemic equilibrium *ξ_E_*, of the syphilis-only sub-model as *t* → *∞* for 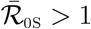.

The epidemiological implication of Theorem 3.3 is that, if a previous infection with syphilis confers lifetime protection against re-infection, then syphilis infection will persist in the population, if the threshold quantity, 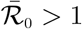.

### 3.2 HPV-Only sub-model

The HPV-only sub-model is (obtained by setting *E* = *I* = *L* = *T* = *H*_E_ = *H*_I_ = *H*_L_ = *P*_E_ = *P*_I_ = *P*_L_ = *C*_E_ = *C*_I_ = *C*_L_ = 0 in the model (1)) given by:

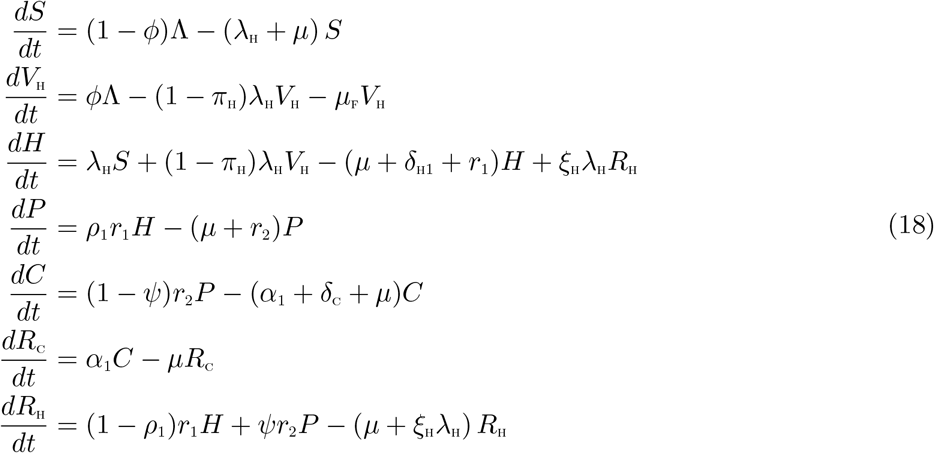

where now,

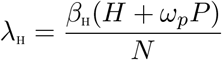

with

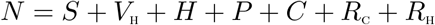

#### 3.2.1 Basic reproduction number of the HPV-only sub-model

The HPV-only sub-model (18) has a DFE, obtained by setting the right-hand sides of the equations in the model (18) to zero, given by

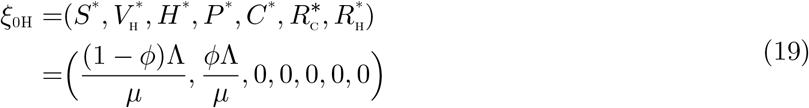

The basic reproduction number, using the next generation operator method [42] is given by

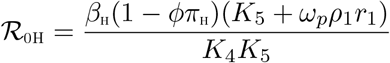

with

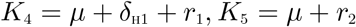

The local and global asymptotic stability analyses of the HPV-only sub-model (18) were well explored by Omame *et al*. [30].

## 4 Local asymptotic stability of disease-free equilibrium co-infection model (1)

**Lemma 4.1** *The DFE, ξ*_0_, *of the HPV-Syphilis co-infection model* (1) *is locally asymptotically stable (LAS) if* 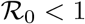, *and unstable if* 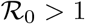.

The epidemiological implication of Lemma 4.1 is that when 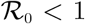, a small influx of HPV or syphilis-infected individuals into the population will not generate large HPV or syphilis outbreaks, and the diseases will die out.

### 4.1 Global asymptotic stability of the disease free equilirium of the full co-infection model (1)

Using the same approach as in Section 3.1.3, we can establish the following result.

Consider the conditions below:

(*H*1): For 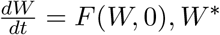 is globally asymptotically stable (GAS),
(*H*2): 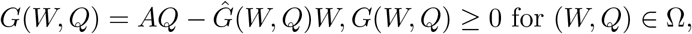

where *A* = *D_Q_G*(*W^∗^*, 0) is an M-matrix (the off-diagonal elements of *A* are nonnegative) and Ω is the region where the model makes biological sense. If System 1 satisfies the above two conditions then the following theorem holds:

**Theorem 4.1** *The fixed point U*_0_ = (*W^∗^*, 0) *is a globally asymptotic stable (GAS) equilibrium of 1 provided that R*_0_ < 1 *(LAS) and that assumptions* (*H*1) *and* (*H*2) *are satisfied*

The Proof follows as in section 3.1, where

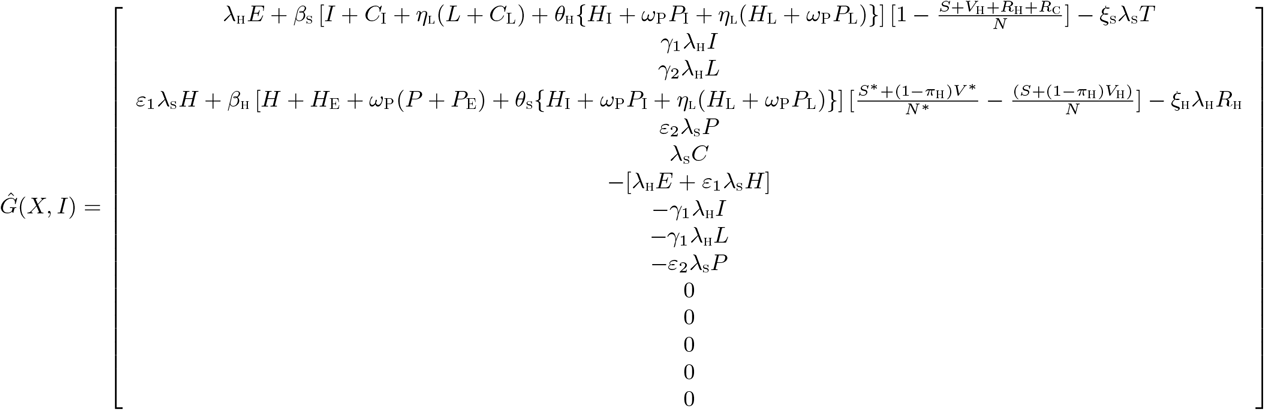

It is evident from the above, that, 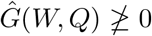, which means that the condition (*H*2) is not satisfied. Hence the DFE, *U*_0_ = (*W^∗^*, 0) may not be globally asymptotically stable, suggesting the possibility of a backward bifurcation. This supports the backward bifurcation analysis for the full-co-infection model (1) in the next section.

### 4.2 Backward bifurcation analysis of the full co-infection model (1)

We shall investigate the type of bifurcation the model (1) may undergo, using the Centre Manifold Theory as discussed in [10]. The following result can be obtained using the approach in [10].

**Theorem 4.2** *If* 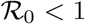 *and a backward bifurcation coefficient a >* 0, *where*

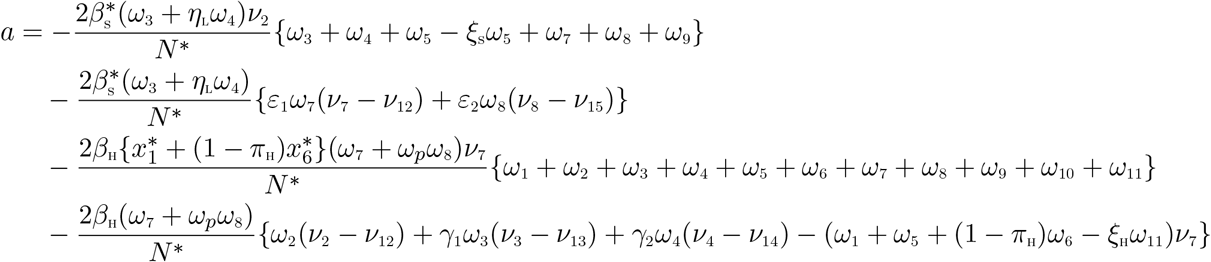

*then model* (1) *exhibits backward bifurcation at* 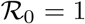. *If a* < 0, *then the system* (1) *exhibits a forward bifurcation at R*_0_ = 1.

**Proof** Suppose

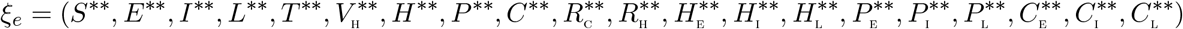

represents any arbitrary endemic equilibrium of the model (that is, an endemic equilibrium in which at least one of the infected components is non-zero). To apply the Centre Manifold Theory, it is necessary to carry out the following change of variables.

Let

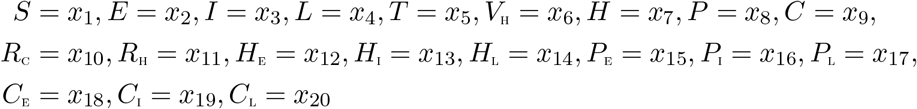

so that

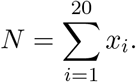

Further, using the vector notation

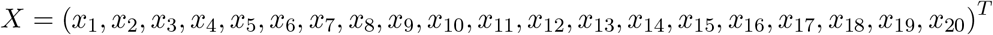

the model (1) can be re-written in the form

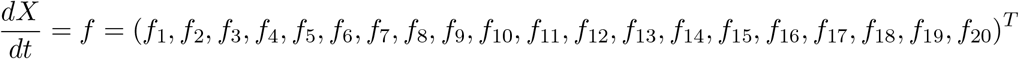

as follows:

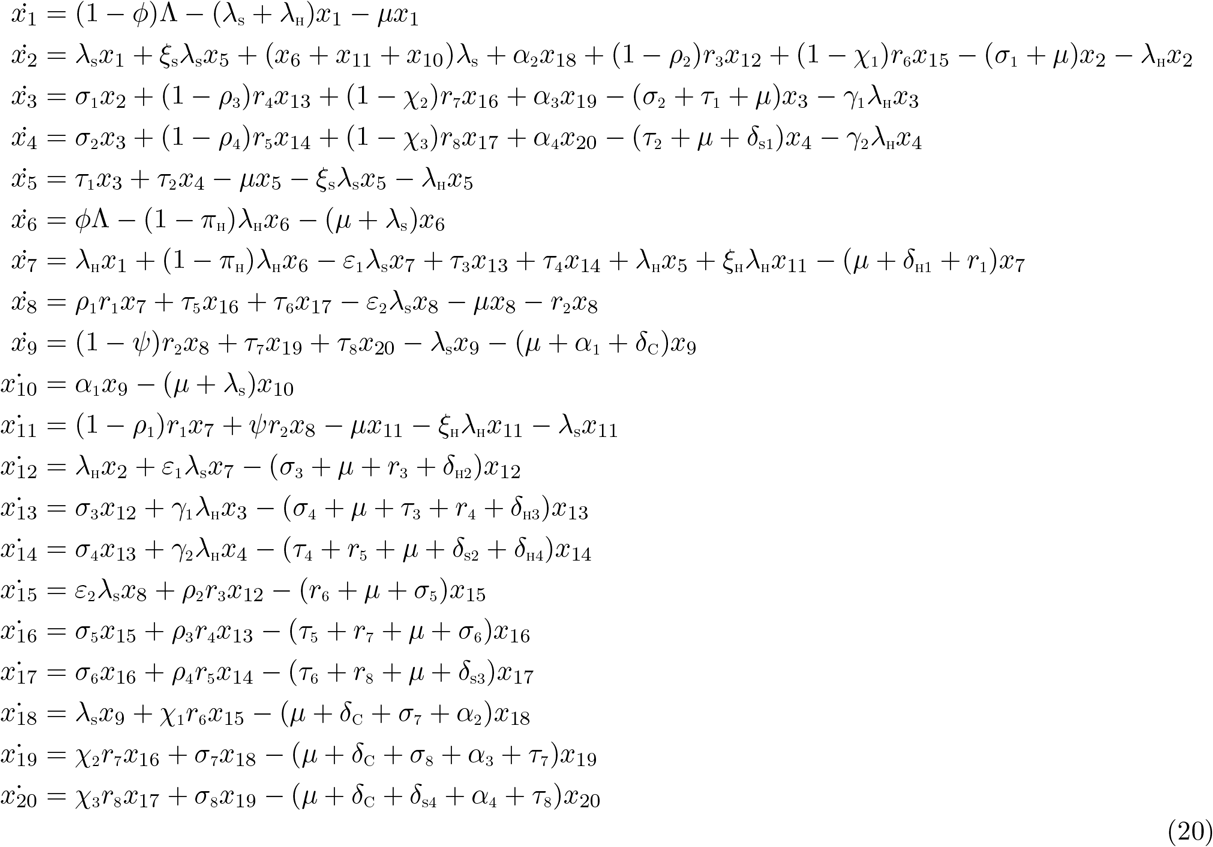

where,

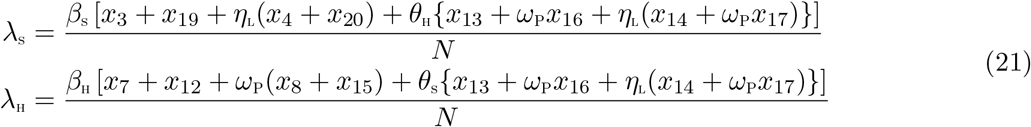

Consider the case when 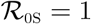. Suppose, further, that *β*_s_ is chosen as a bifurcation parameter. Solving for 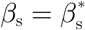 from 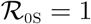 gives

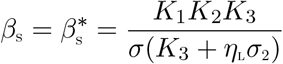

Evaluating the Jacobian of the system (20) at the DFE, *J*(*ξ*_0_), and using the approach in [10], we have that *J*(*ξ*_0_) has a right eigenvector (associated with the simple zero eigenvalue of *J*(*ξ*_0_)) given by

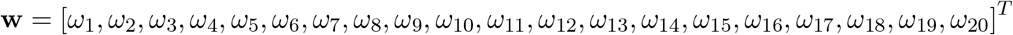

where,

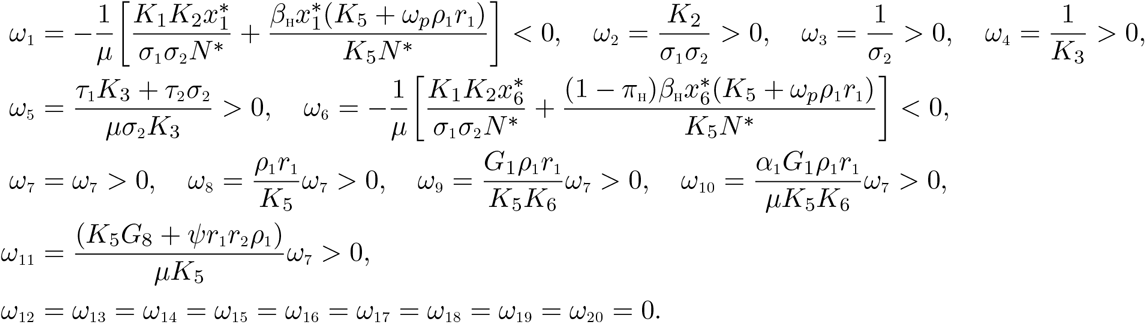

The components of the left eigenvector of 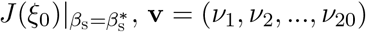, satisfying **v.w** = 1 are

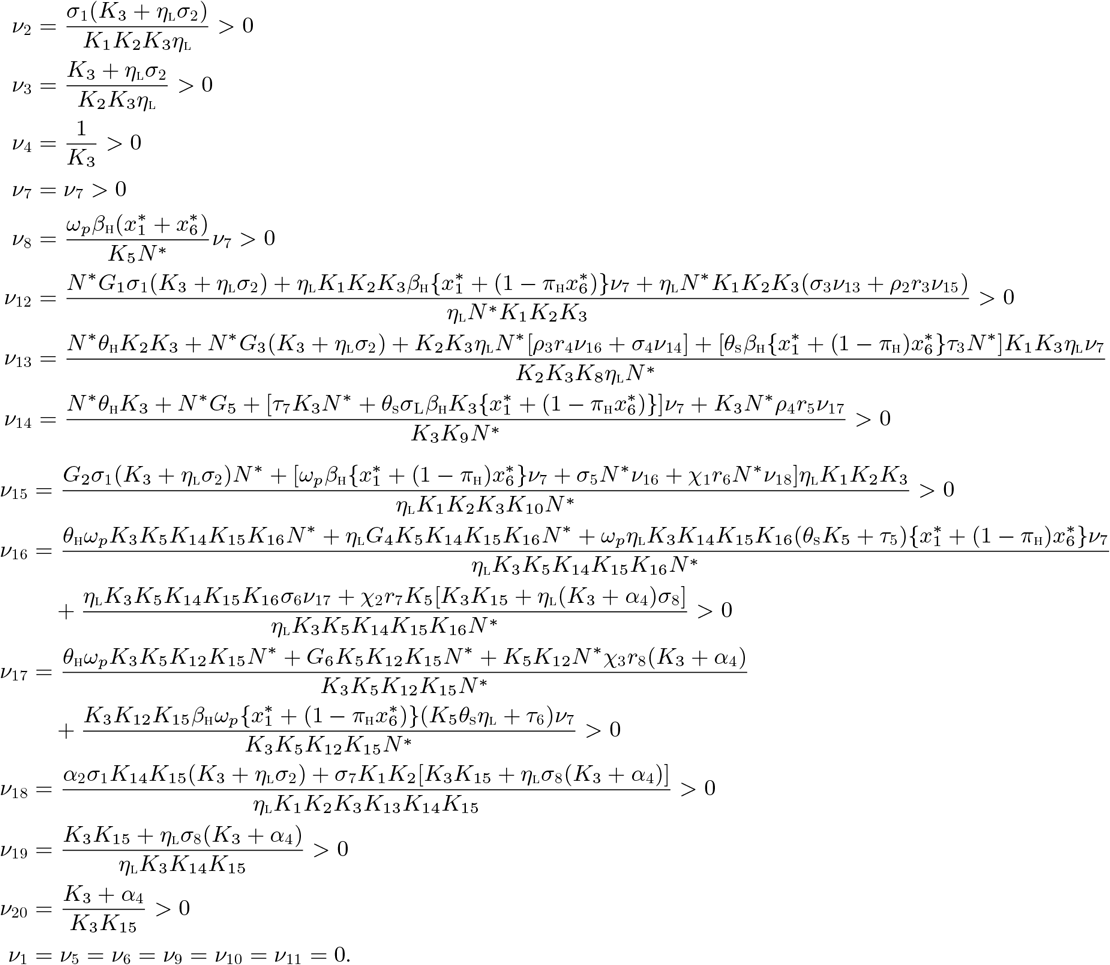

where,

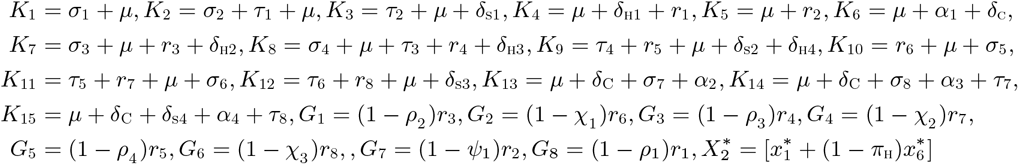

It follows from Theorem 4.1 in [10], by computing the non-zero partial derivatives of *F* (*x*) (evaluated at the disease free equilibrium, (*ξ*_0_)) that the associated bifurcation coefficients defined by *a* and *b*, given by

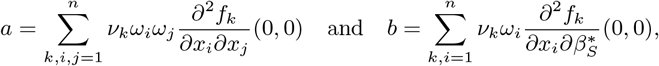

are computed to be

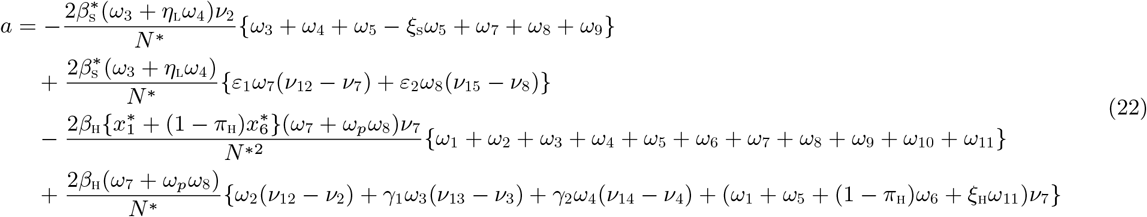

and

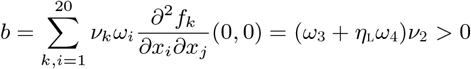

Since the bifurcation coefficient *b* is positive, it follows from Theorem 4.1 in [10] that the model (1), or the transformed model (20), will undergo a backward bifurcation if the backward bifurcation coefficient, *a*, given by (22) is positive.

## 5 Analysis of optimal Control Problem

We apply Pontryagin’s Maximum Principle, in this section, to determine the necessary conditions for the optimal control of the HPV-Syphilis co-infection model. We assume that the proportion of vaccinated individuals, *ϕ* for HPV and the Syphilis treatment rates *u*_1_, *u*_2_, *u*_3_, *u*_4_, *u*_5_, *u*_6_, *u*_7_, *u*_8_ are now time dependent and will therefore act as the control variables. Hence we have,

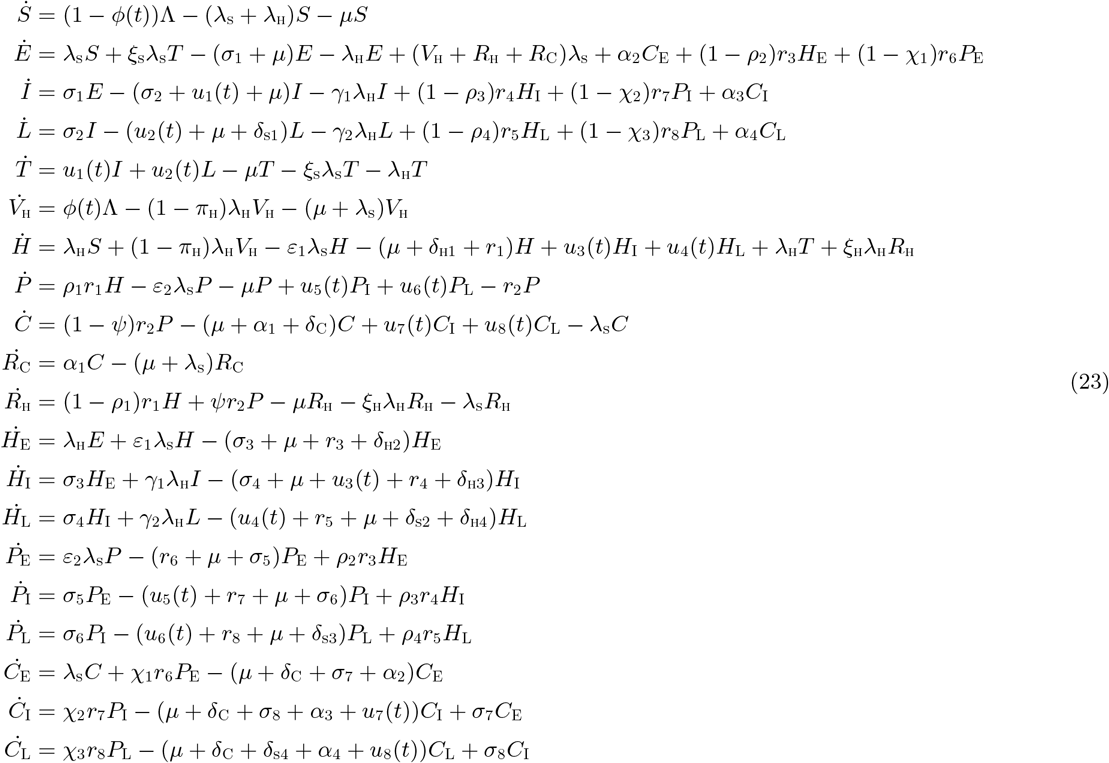

where,

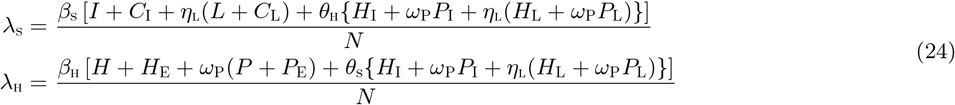

For this, we consider the objective functional

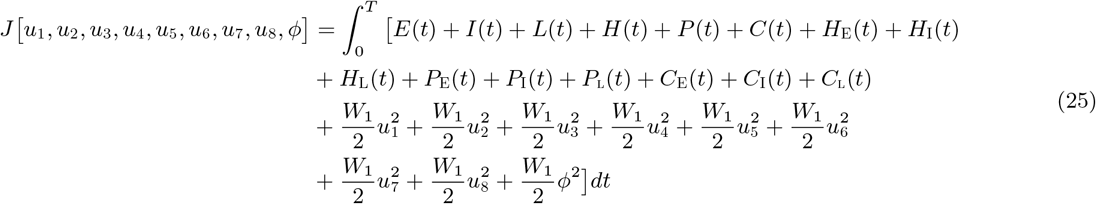

The parameters, *W*_1_ is the weight on the benefit and cost of implementing the optimal control measure, where *W*_1_ balances the cost factors as a result of the size and the relevance of the terms making up the objective functional. *T* is the final time. We seek to find an optimal control, 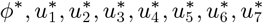 such that

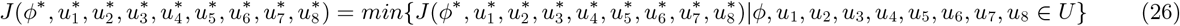

where 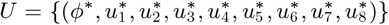 such that 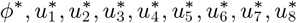 are measurable with 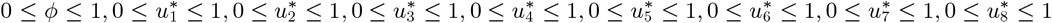, for *t* ∈ [0, *T*] is the control set.

The Pontryagin’s Maximum Principle [33] gives the necessary conditions which an optimal control pair must satisfy.

This principle transforms (23), (25) and (26) into a problem of minimizing a Hamiltonian, 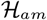, pointwisely with regards to the control functions, *ϕ, u*_1_, *u*_2_, *u*_3_, *u*_4_, *u*_5_, *u*_6_, *u*_7_ and *u*_8_:

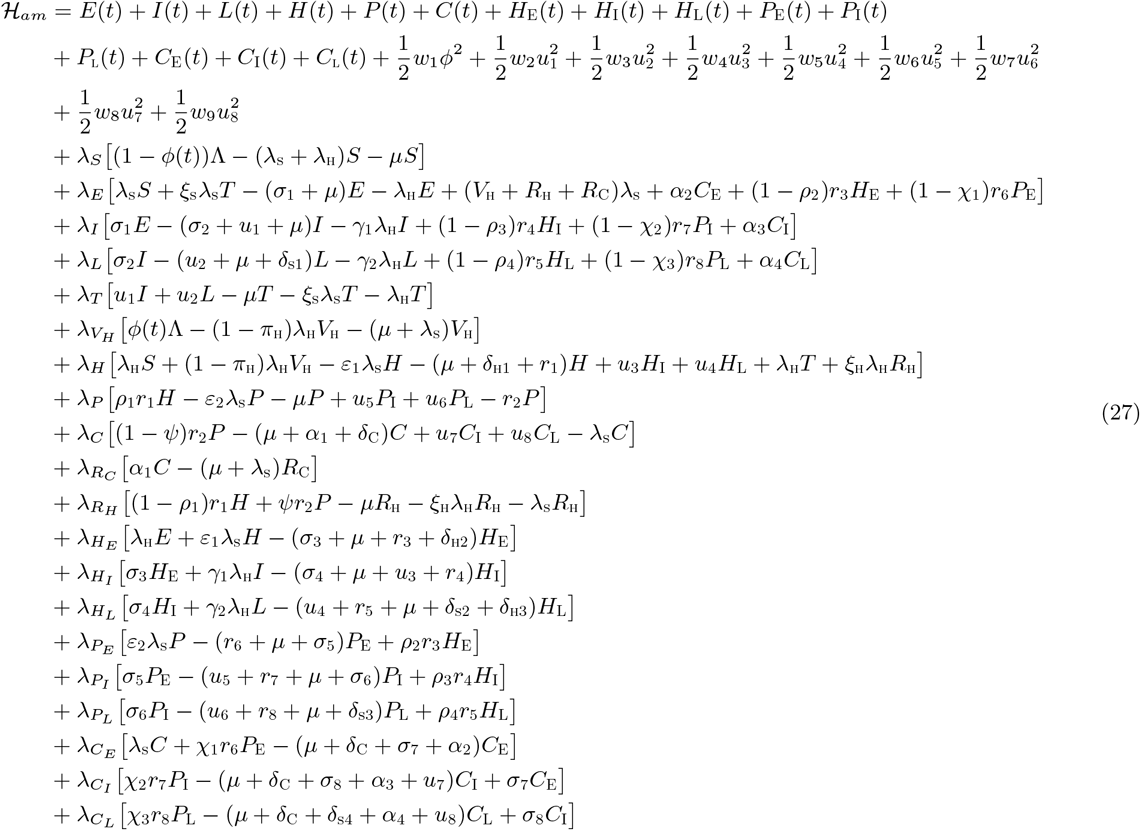

**Theorem 5.1** *For an optimal control set ϕ, u*_1_, *u*_2_, *u*_3_, *u*_4_, *u*_5_, *u*_6_, *u*_7_, *u*_8_ *that minimizes J over U, there are adjoint variables, λ*_1_, *λ*_2_,…, *λ*_20_ *satisfying*

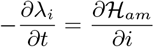

*and with transversality conditions*

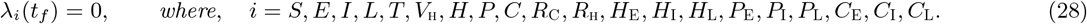

*Furthermore*,

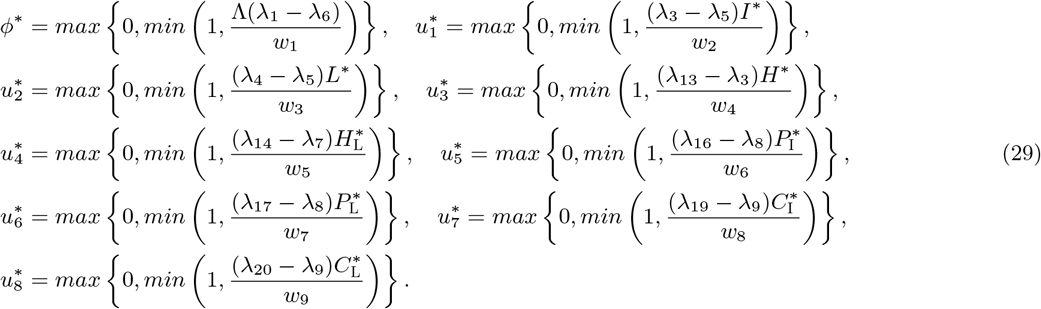

Proof of Theorem 5.1

Suppose 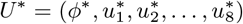 is an optimal control and 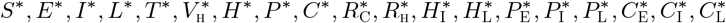 are the corresponding state solutions. Applying the Pontryagin’s Maximum Principle [33], there exist adjoint variables satisfying:

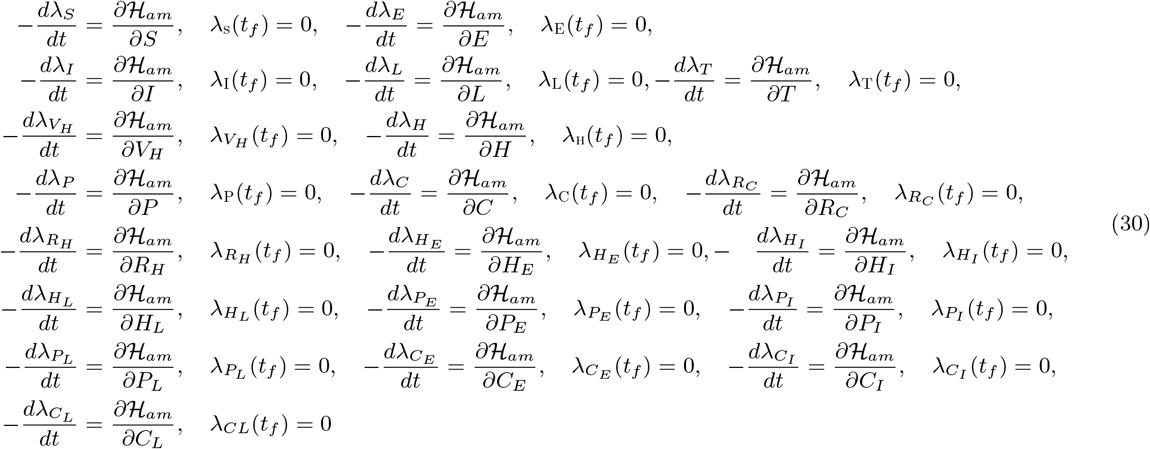

with transversality conditions; *λ_S_* (*t_f_*) = *λ_E_*(*t_f_*) = *λ_I_*(*t_f_*), *λ_L_*(*t_f_*) = *λ_T_* (*t_f_*) = *λ_VH_* (*t_f_*) = *λ_H_*(*t_f_*) = *λ_P_* (*t_f_*) = *λ_C_*(*t_f_*) = *λ_RC_* (*t_f_*) = *λ_RH_* (*t_f_*) = *λ_HE_* (*t_f_*) = *λ_HI_* (*t_f_*) = *λ_HL_*(*t_f_*) = *λ_PE_* (*t_f_*) = *λ_PE_* (*t_f_*) = *λ_PI_* (*t_f_*) = *λ_PL_*(*t_f_*) = *λ_CE_* (*t_f_*) = *λ_CI_* (*t_f_*) = *λ_CL_*(*t_f_*) = 0 We can determine the behaviour of the control by differentiating the Hamiltonian, 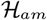 with respect to the controls(*ϕ, u*_1_, *u*_2_, *u*_3_, *u*_4_, *u*_5_, *u*_6_, *u*_7_, *u*_8_) at *t*. On the interior of the control set, where 0 *< u_j_* < 1 for all (*j* = 1,…, 8) and 0 *< ϕ* < 1, we obtain

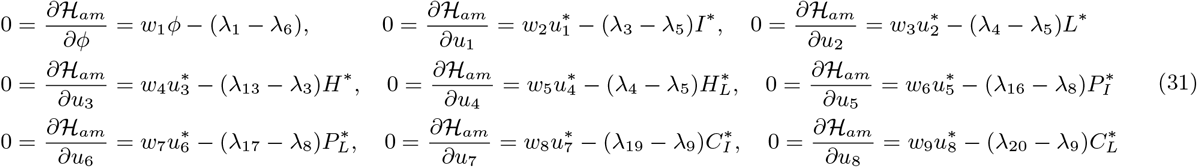

Therefore, we have that [20]

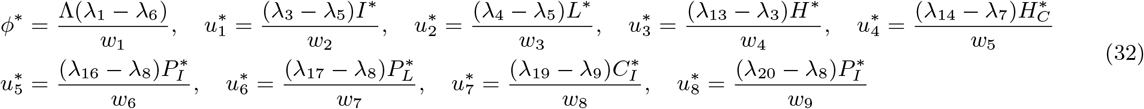

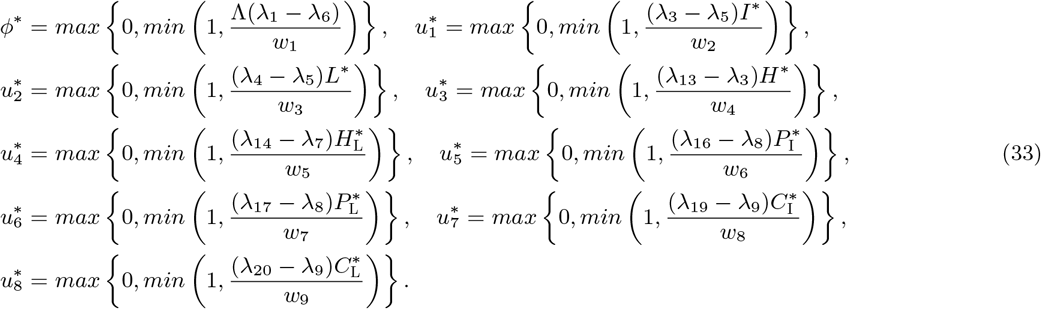

### 5.1 Numerical simulations

We now simulate the optimal control model (23) numerically using the parameter estimates in Table 2, so that the reproduction number, 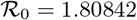 (unless otherwise stated), to assess the potential impact of various targeted control strategies on the transmission dynamics of HPV and SYphilis in the population. Demographic parameters relevant to the city of Rio de Janeiro in Brazil were chosen. Specifically, since the total population of sexually active susceptible individuals (15-64 years) in the State of Rio de Janeiro in Brazil are estimated to be 12,850,804, at disease-free equilibrium, 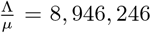[9]. In Brazil, the life expectancy is estimated at 74 years [9]. Hence we have that *µ* = 0.0135, so that Λ = 120895 per year.

**Table 2:** Description of parameters in the model (1).

| Parameter | Description | Value | References |
| --- | --- | --- | --- |
| $\Lambda_H$ | Recruitment rate for humans | 120895 | [9] |
| $\mu_H$ | Natural death rate | 0.0135 | [9] |
| $\phi$ | fraction of individuals vaccinated against HPV | 0.70 | Assumed |
| $\pi_H$ | HPV vaccine efficacy rate | 0.90 | [29] |
| $\tau_1, \tau_2$ | Syphilis treatment rate for singly infected in compartments $I$ and $L$, respectively | 3.65 | Implied from [16] |
| $r_1, r_2$ | HPV natural recovery or clearance rate for individuals in compartments $H$ and $P$, respectively | 0.9 | [30] |
| $\beta_S$ | Effective contact rate for syphilis infection | 7.0 | [23] |
| $\beta_H$ | Effective contact rate for HPV infection | 2.0 | [30] |
| $r_i \quad i=3, \dots, 8$ | HPV natural recovery or clearance rates for dually infected individuals in compartments $H_E, H_I, H_L, P_E, P_I$ and $P_L$, respectively | 0.9 | [30] |
| $\tau_i \quad i=3, \dots, 8$ | Syphilis treatment rates for dually infected individuals in compartments $H_I, H_L, P_I, P_L, C_I$ and $C_L$, respectively | 1.0 | [27] |
| $\delta_{s1}$ | Syphilis-induced death rate for singly infected individuals | 0.06849 | [14] |
| $\delta_{s2}, \delta_{s3}, \delta_{s3}$ | Syphilis-induced death rates for dually infected individuals in compartments $H_L, P_L$ and $C_L$, respectively | 0.06849 | Assumed |
| $\delta_{H1}$ | HPV-induced death rate for singly infected individuals | 0.001 | [29] |
| $\delta_C$ | Cancer-induced death rate | 0.001 | [31] |
| $\delta_{H2}, \delta_{H3}, \delta_{H4}$ | HPV-induced death rates for dually infected individuals in compartments $H_E, H_I$ and $H_L$, respectively | 0.001 | [30] |
| $\alpha_i, \quad i=1, 2, 3, 4$ | Anal cancer treatment rates for individuals in compartments $C, C_E, C_I$ and $C_L$, respectively | 0.76 | [21] |
| $\eta_L$ | Modification parameter accounting for infectiousness of syphilis infected individuals in late latent stage | 0.001 | [21] |
| $\theta_H$ | Modification parameter accounting for increased infectiousness of dually infected individuals due to HPV infection | 1.3 | Assumed |
| $\theta_S$ | Modification parameter accounting for increased infectiousness of dually infected individuals due to syphilis infection | 1.3 | Assumed |
| $\xi_S$ | Syphilis re-infection rate | 0.6 | [24] |
| $\xi_H$ | HPV re-infection rate | 0.2 | [31] |
| $\omega_P$ | Modification parameter accounting for infectiousness of persistent HPV-infected individuals | 0.9 | [4] |
| $\omega_P$ | Modification parameter accounting for infectiousness of persistent HPV-infected individuals | 0.9 | [4] |
| $\psi$ | fraction of persistent HPV-infected individuals who recover naturally and do not develop anal cancer | 0.5 | [21] |
| $\gamma_i \quad i=1, 2, 3$ | Modification parameter accounting for the susceptibility of syphilis-infected individuals to HPV infection | 1.2 | [34, 35] |
| $\varepsilon_i \quad i=1, 2, 3$ | Modification parameter accounting for the susceptibility of HPV-infected individuals to syphilis infection | 1.3 | [26, 25] |
| $\rho_i \quad i=1, 2, 3$ | Fraction of HPV-infected individuals who develop persistent HPV infection | 0.5 | Assumed |
| $\sigma_1, \sigma_3, \sigma_5$ | Progression rates to early stage of syphilis infection | 0.2 | Assumed |
| $\sigma_2, \sigma_4, \sigma_6$ | progression rates to late stage of syphilis infection | 0.2 | [16] |

Numerical simulations of the optimal control problem (23), adjoint equations (30) and characterizations of the control (33) are implemented by the Runge Kutta method using the forward backward sweep (carried out in MATLAB). The balancing factor *w*_1_ = 10^3^ (The Centres for Disease Control cost per dose of the pediatric *Gardasil 4*, the *Gardasil 9*, and the *Cervarix*, respectively, is $121.03,$134.26 and $107.97 [12, 22]. Since the associated costs considered are combinations of price per dose, storage/ administration-related costs and transportation, etc, we will assume *w*_1_ = 10^3^). Following the estimates for syphilis treatment in [18], the balancing factors for syphilis-infected individuals are: *w*_2_ = *w*_3_ = 200, *w*_4_ = *w*_5_ = *w*_6_ = *w*_7_ = *w*_8_ = *w*_9_ = 220. Here, we assume that the cost of treatment for co-infected individuals is more than the cost of treatment for singly-infected individuals. Following the reported prevalence of Syphilis and HPV as well as the co-infection prevalence in [34, 39] and the demographic data obtained from [9], we set the initial conditions at: *S*(0) = 100, 000, *E*(0) = 10, 000, *I*(0) = 20, 000, *L*(0) = 2000, *T* (0) = 0, *V_H_* = 100, 000, *H*(0) = 2000, *P* (0) = 1000, *C*(0) = 2000, *R_C_*(0) = 0, *R_H_*(0) = 0, *H_E_*(0) = 2000, *H_I_*(0) = 10, 000, *H_L_*(0) = 10, 000, *P_E_*(0) = 1000, *P_I_*(0) = 1000, *P_L_*(0) = 1000, *C_E_*(0) = 1000, *C_I_*(0) = 10, 000, *C_L_*(0) = 5000. We implement the following four different control strategies for numerical simulations of the co-infection model 23.

I. Optimal HPV vaccination strategy (*ϕ* ≠ 0);
II. Syphilis treatment controls for singly infected individuals (*u*_1_ ≠ *u*_2_ ≠ 0);
III. Syphilis treatment controls for dually infected individuals only (*u*_3_ ≠ *u*_4_ ≠ *u*_5_ ≠ *u*_6_ ≠ *u*_7_ ≠ *u*_8_ ≠ 0);
IV. Universal strategy (*ϕ* ≠ *u*_1_ ≠ *u*_2_ ≠ *u*_3_ ≠ *u*_4_ ≠ *u*_5_ ≠ *u*_6_ ≠ *u*_7_ ≠ *u*_8_ ≠ 0).

The control profiles for each of the various controls are presented in Figures 8, 10 and 12.

#### 5.1.1 Strategy I: Optimal HPV vaccination strategy (*ϕ* ≠ 0)

Applying this strategy, we note in Figures 3 (a) and 3(b) that the total number of individuals singly infected with HPV and persistent HPV, respectively, is less than the total number when no control is applied. It can equally be observed that optimal HPV vaccination strategy has a high positive population level impact on the populations of infected individuals dually infected with HPV and Syphilis in early and late stages of infection, respectively, (Figures 4 (a) and 4(b)), infected individuals dually infected with persistent HPV and Syphilis in early and late stages of infection,respectively (Figure 5(a) and 5(b)) and infected individuals dually infected with Cancer and Syphilis in early and late stages of infection, respectively(Figure 6 (a) and 6(b)). The results of the simulations agree with the epidemiological reports in [26, 25], that prior HPV infections are risk factors for syphilis infection. Therefore, controlling HPV infections through vaccination can bring down the burden of co-infection of the two diseases.

**Figure 3:**
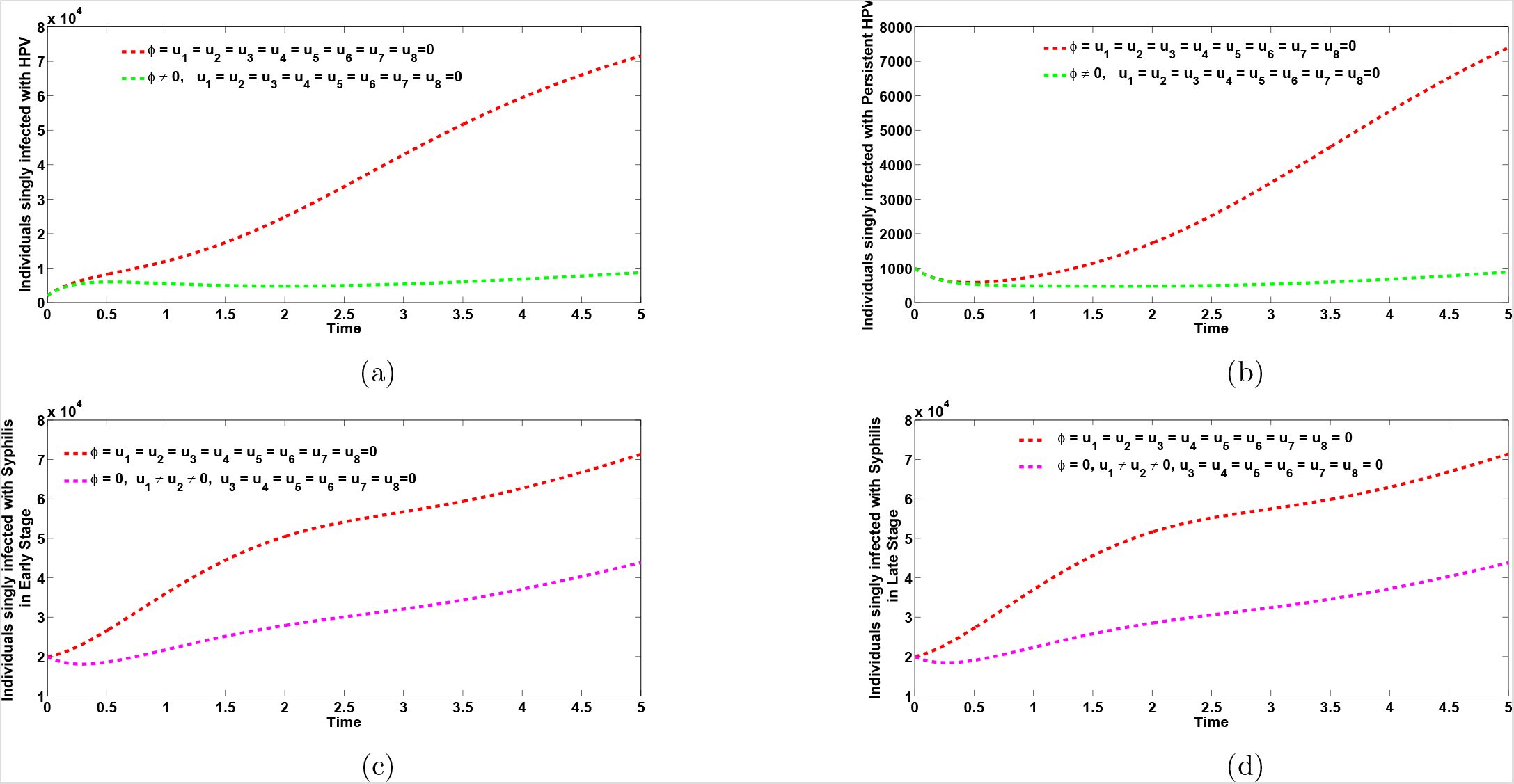
Plots of the total number of individuals singly infected with HPV and persistent HPV, respectively (Figures 3a and 3b) in the presence of optimal vaccination control only (*ϕ* ≠ 0) and total number of individuals singly infected with syphilis in early and late stages of infection, respectively (Figures 3c and 3d) in the presence of treatment controls for singly infected only (*u*_1_ ≠ *u*_2_ ≠ 0). Here, *β*_s_ = 7.0, *β*_H_ = 2.0. All other parameters as in Table 2

**Figure 4:**
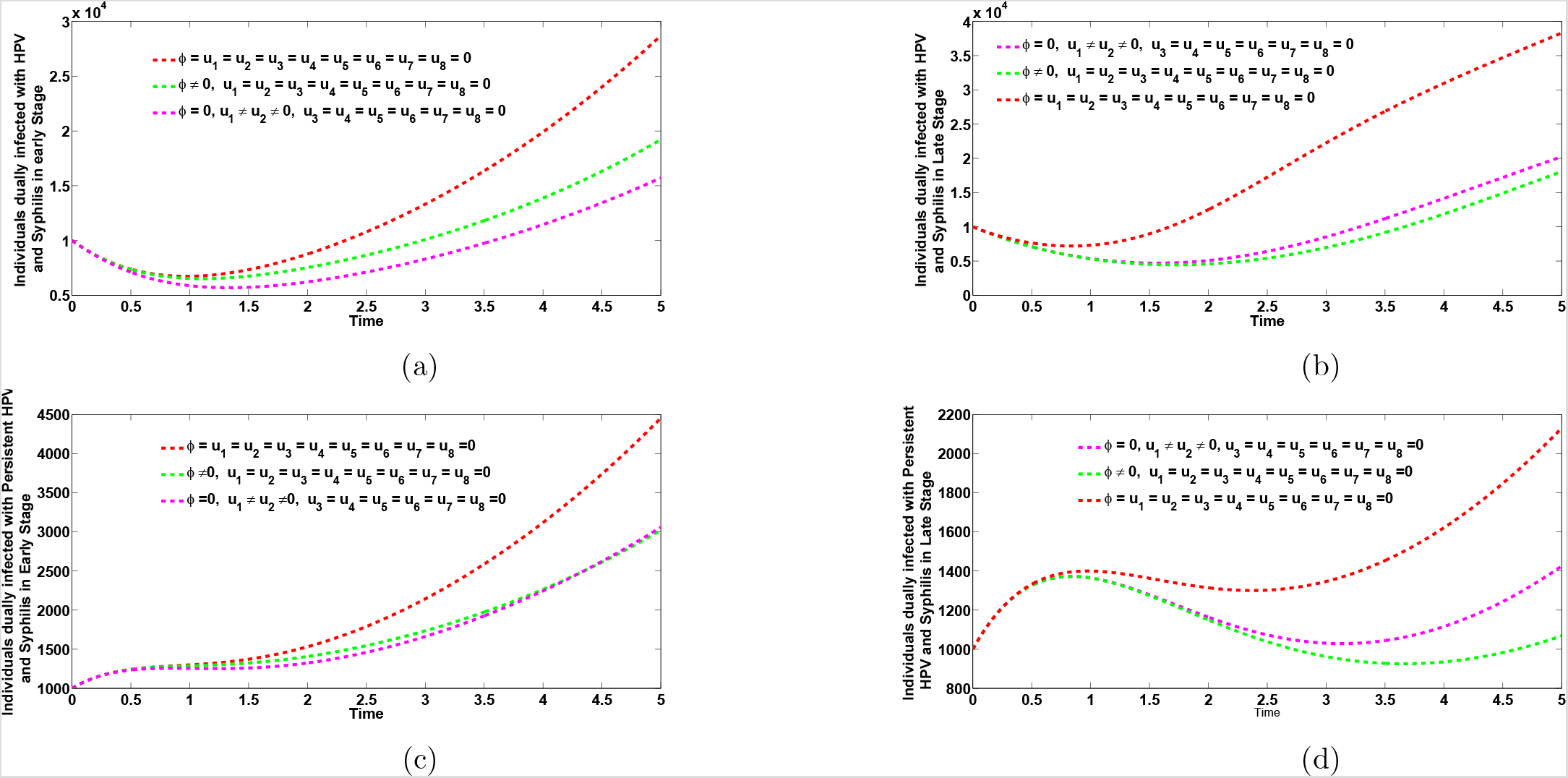
Plots of the co-infection cases for individuals dually infected with HPV and syphilis in early and late stages of infection, respectively (Figure 4a and 4b) and co-infection cases for individuals dually infected with persistent HPV and syphilis in early and late stages of infection, respectively (Figures 4c and 4d), when optimal vaccination control only strategy (*ϕ* ≠ 0) and syphilis treatment controls for singly-infected (*u*_1_ ≠ *u*_2_ ≠ 0) are administered. Here, *β*_s_ = 7.0, *β*_H_ = 2.0. All other parameters as in Table 2

**Figure 5:**
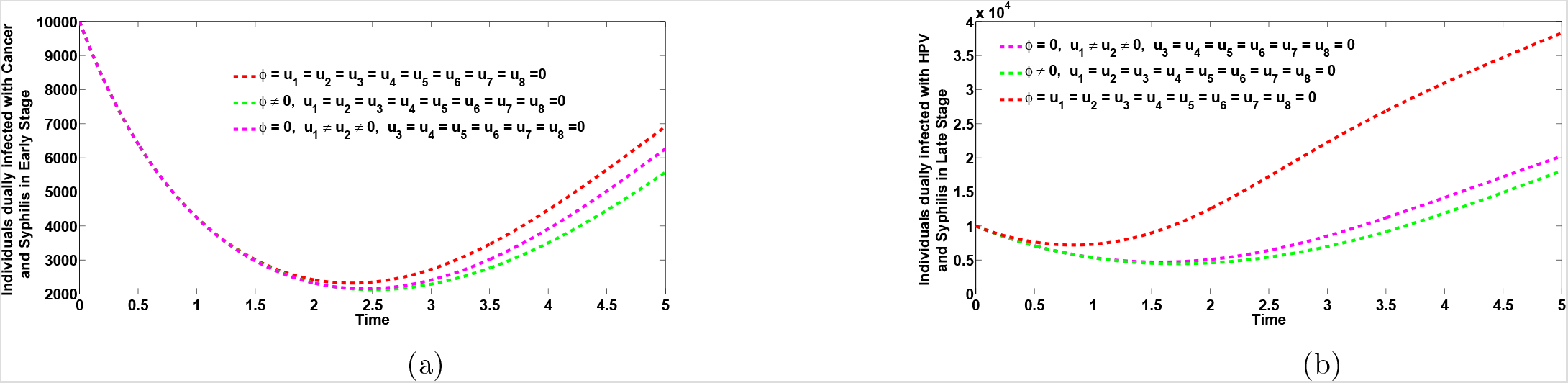
Plots of the co-infection cases for individuals dually infected with anal cancer and syphilis in early and late stages of infection, respectively (Figure 5a and 5b), when optimal vaccination control only strategy (*ϕ* ≠ 0) and syphilis treatment controls for singly-infected (*u*_1_ ≠ *u*_2_ ≠ 0) are administered. Here, *β*_s_ = 7.0, *β*_H_ = 2.0. All other parameters as in Table 2

**Figure 6:**
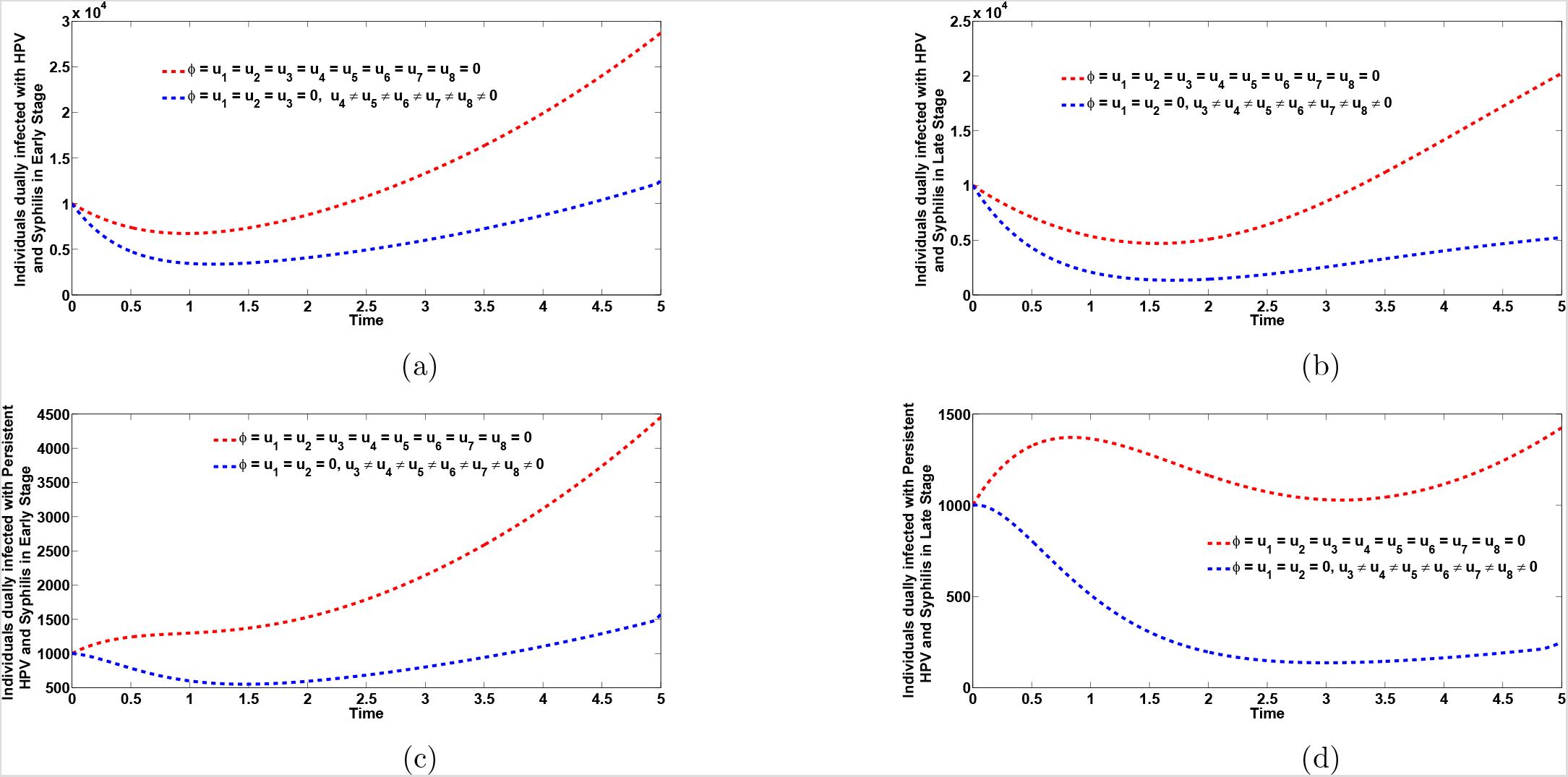
Plots of the co-infection cases for individuals dually infected with HPV and syphilis in early and late stages of infection, respectively (Figures 6(a) and 6b) as well as co-infection cases for individuals dually infected with persistent HPV and syphilis in early and late stages of infection, respectively (Figures 6c and 6d). Here, *β*_s_ = 7.0, *β*_H_ = 2.0, *ϕ* = *u*_1_ = *u*_2_ = 0, *u*_3_ ≠ *u*_4_ ≠ *u*_5_ ≠ *u*_6_ ≠ *u*_7_ ≠ *u*_8_ ≠ 0. All other parameters as in Table 2

**Figure 7 Figure 8:**
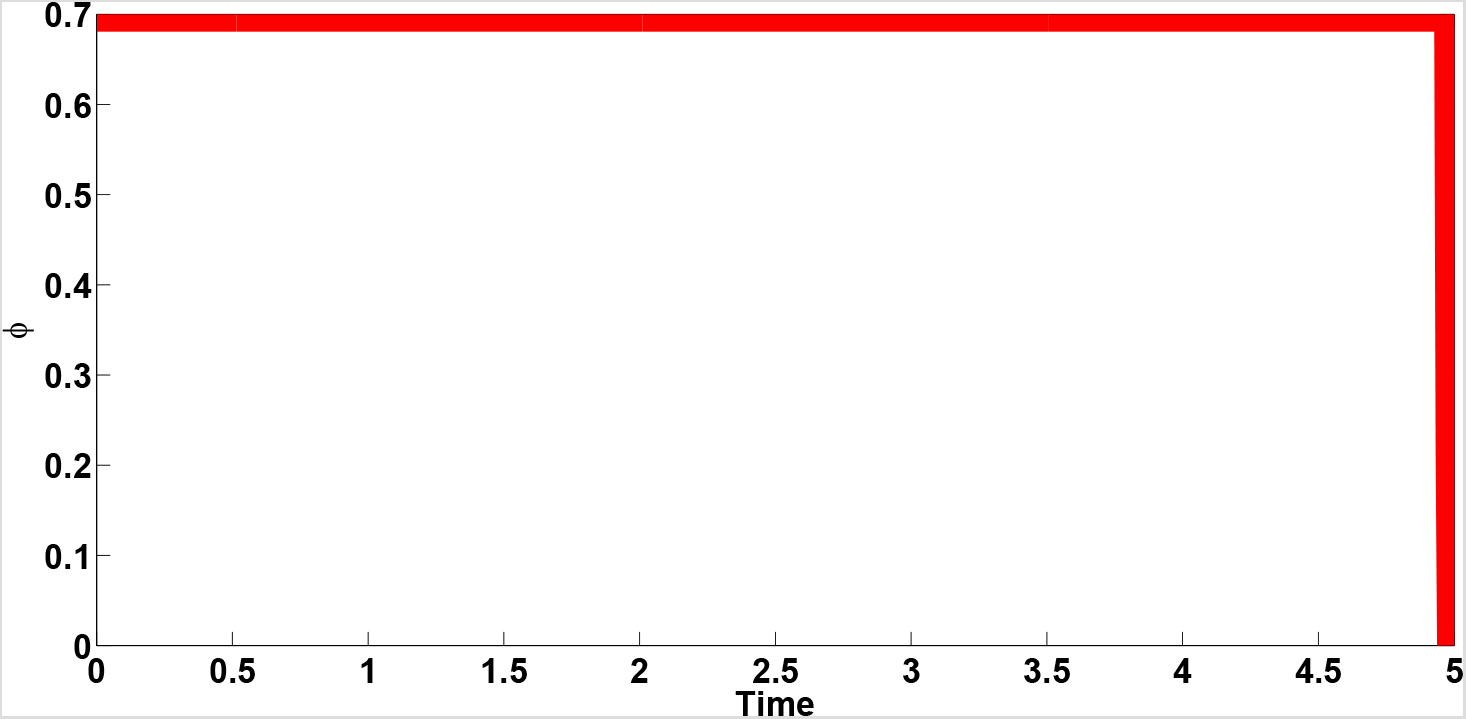
Effects of optimal control *ϕ* on the dynamics of the co-infection optimal control model (23)

**Figure 9 Figure 10:**
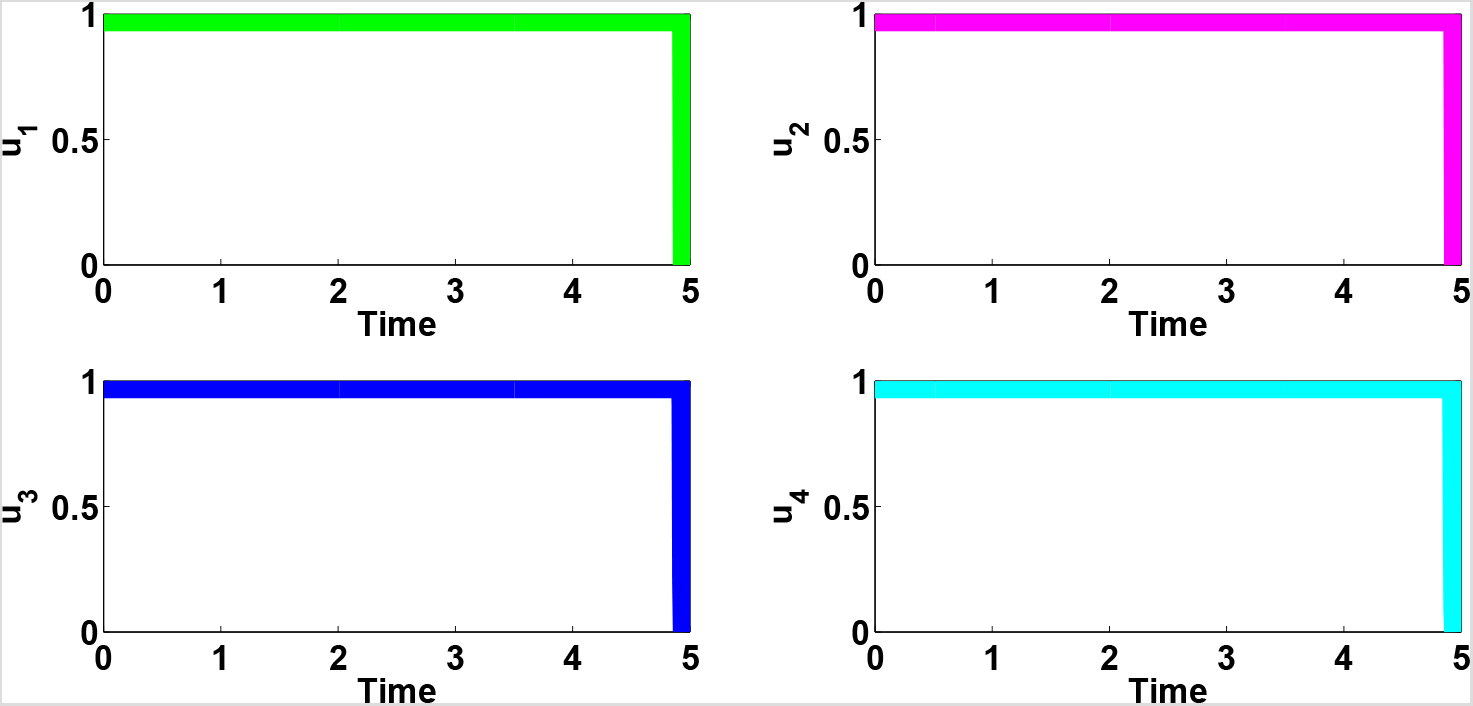
Combined effects of optimal controls *u*_1_, *u*_2_, *u*_3_ and *u*_4_ on the dynamics of the co-infection optimal control model (23)

**Figure 11 Figure 12:**
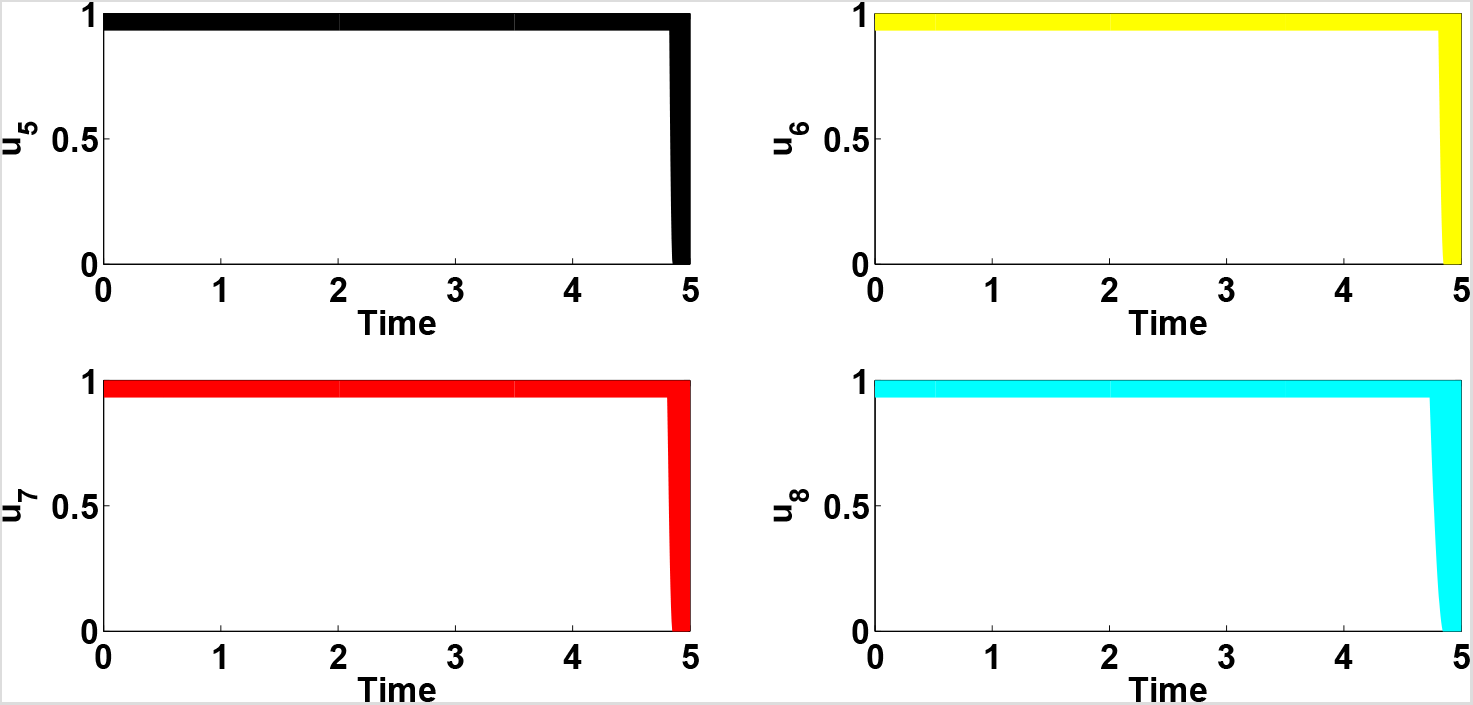
Combined effects of optimal controls *u*_5_, *u*_6_, *u*_7_ and *u*_8_ on the dynamics of the co-infection optimal control model (23)

#### 5.1.2 Strategy II: Syphilis treatment controls for singly infected individuals only (*u*_1_ *6* ≠ *u*_2_ ≠ 0)

Using this control strategy, we observe in Figure (3(c) and 3 (d), that the number of individuals singly infected with Syphilis in early and late stages of infection, respectively, is less than the number when no control strategy is apllied. Likewise, the Syphilis only treatment controls for singly infected with syphilis has positive positive population level impact on the populations of infected individuals dually infected with HPV and Syphilis in early and late stages of infection, respectively, Figures(4 (c) and 4 (d). Treatment controls for singly infected with syphilis equally has positive population level impact on the population of individuals dually infected with persistent HPV and Syphilis in early and late stages of infection as observed in Figure (5 (c) and 5 (d). The total number of individuals dually infected with Cancer and Syphilis in early and late stages of infection is lesser when treatment controls are applied to individuals singly infected with Syphilis than when no treatment control is applied (Figures 6 (c) and 6 (d)).

#### 5.1.3 Strategy III: Syphilis treatment controls for dually infected individuals only (*u*_3_ *6* ≠ *u*_4_ ≠ *u*_5_ ≠ *u*_6_ ≠ *u*_7_ ≠ *u*_8_ ≠ 0)

The simulations of the total number of dually infected individuals in the presence of Syphilis treatment controls are depicted in Figures 6a–6d). Applying this control, we observe that the total number of individuals dually infected with HPV and Syphilis in early and late stages of infection respectively, is less than the total population when no control is applied as expected (Figure 6(a) and 6(b)). Similarly, it is noticed from Figures (6 (c) and 6 (d), that the total number of individuals dually infected with persistent HPV and Syphilis in early and late stages of infection, respectively, is lesser when this control is applied. In addition high population level impact is noticed on the total number of individuals dually infected with Cancer and Syphilis in early and late stages of infection, respectively, as shown in (Figure 13(a) and 13 (b). This supports the epidemiological report in the introduction section that syphilis is a risk factor for HPV infection [34]. Hence, if we focus on syphilis treatment controls, it can significantly bring down the burden of the co-infection of HPV and syphilis in a population. The simulations equally agree with the findings in Tseng *et al*. [35], that prior syphilis infection was associated with persistent HPV and increased susceptibility to anal cancer. As a result, treating syphilis infection in individuals dually infected with persistent HPV and anal cancer or focusing on syphilis treatment among individuals dually infected with syphilis and anal cancer, will significantly curb the mixed infections.

#### 5.1.4 Strategy IV: Universal strategy (*ϕ* ≠ *u*_1_ ≠ *u*_2_ ≠ *u*_3_ ≠ *u*_4_ ≠ *u*_5_ ≠ *u*_6_ ≠ *u*_7_ ≠ *u*_8_ ≠ 0)

The simulations of the optimal control model (23) in the presence of combined optimal HPV vaccination strategy and syphilis treatment controls are depicted in Figures 14, 15 and 16. It is observed in Figure 14 that the combined control stratgy has a high population level impact on the populations of individuals singly infected with syphilis in early stage (Figure 14 (a)), individuals singly infected with syphilis in late stage (Figure 14 (b)), individuals singly infected with HPV (Figure 14 (c)) and individuals singly infected with persistent HPV (Figure 14 (d)). In a similar manner, the total number of individuals dually infected with HPV and syphilis in early and late stages of infection, respectively (Figures 15 (a) and 15 (b)), total number of individuals dually infected with persistent HPV and syphilis in early and late stages of infection (Figures 15 (c) and 15 (d)) and total number of individuals dually infected with anal cancer and syphilis in early and late stages of infection, respectively (Figures 16 (a) and 16(b)) all recorded lesser population number when the universal strategy is implemented than no no control is administered.

**Figure 13:**
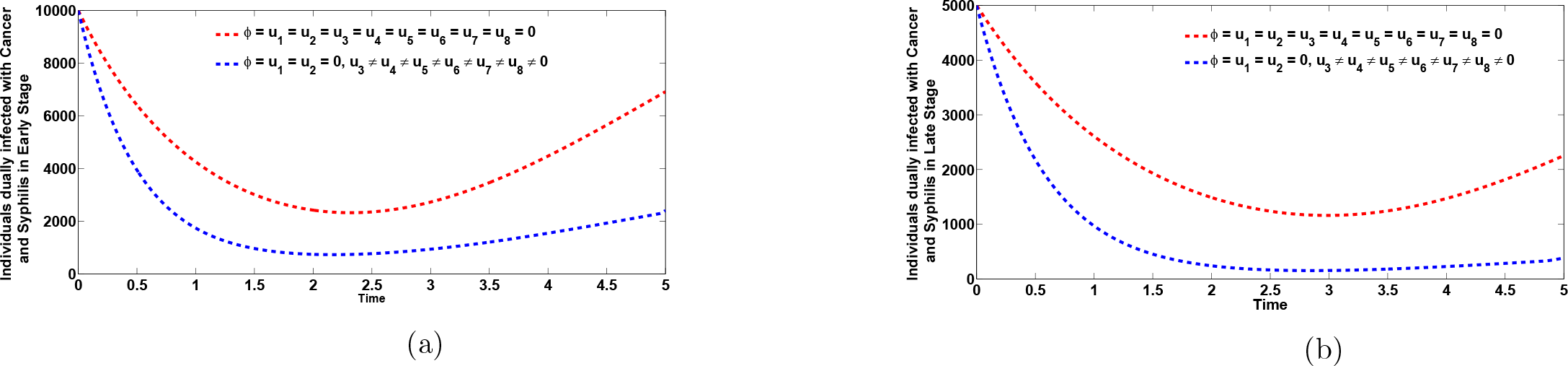
Plots of the co-infection cases for individuals dually infected with anal cancer and syphilis in early and late stages of infection, respectively (Figures 13(a) and 13b). Here, *β*_s_ = 7.0, *β*_H_ = 2.0, *ϕ* = *u*_1_ = *u*_2_ = 0, *u*_3_ ≠ *u*_4_ ≠ *u*_5_ ≠ *u*_6_ ≠ *u*_7_ ≠ *u*_8_ ≠ 0. All other parameters as in Table 2

**Figure 14:**
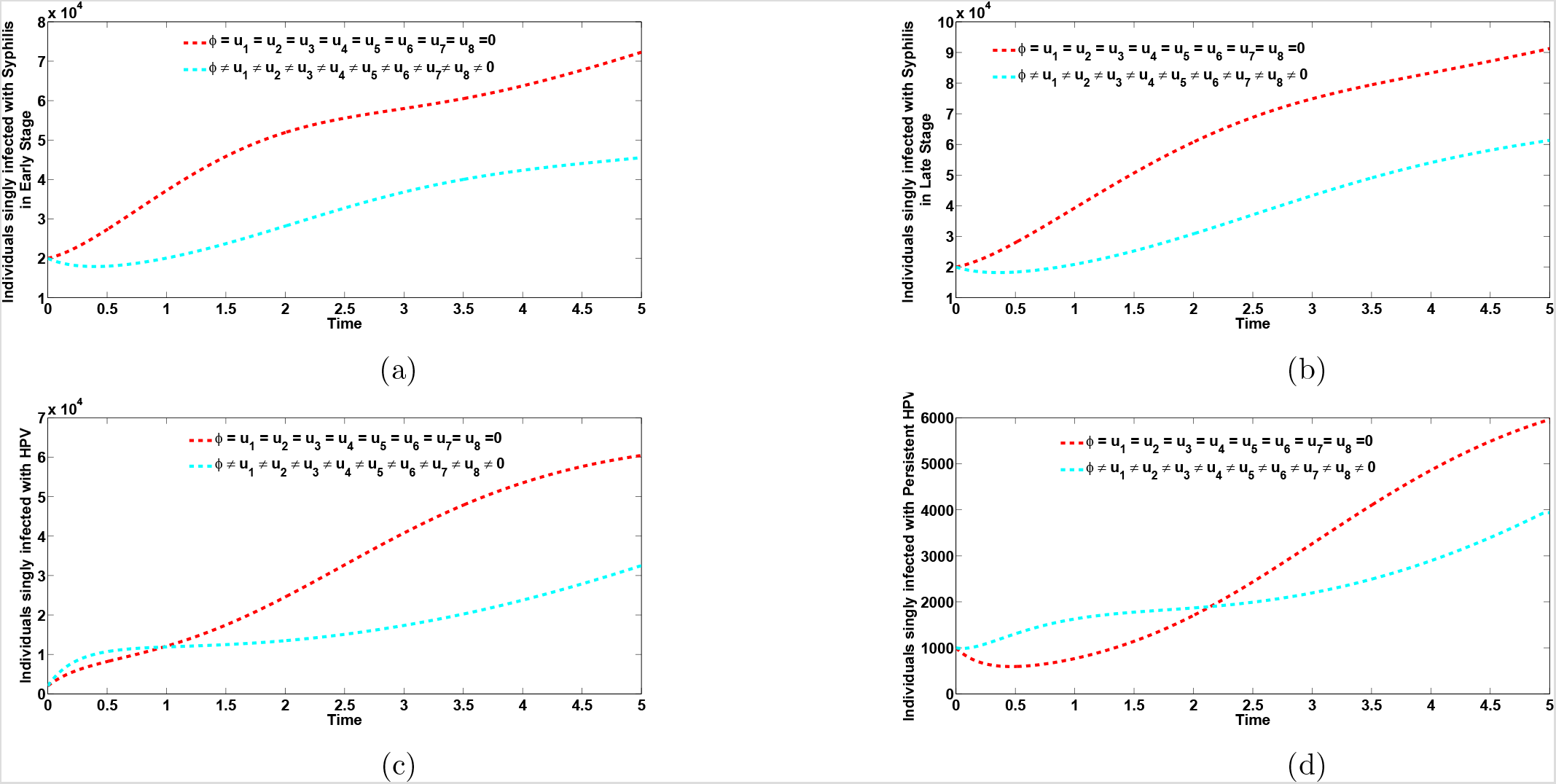
Plots of the total number of infected individuals singly infected with syphilis in early and late stages of infection, respectively (Figures 14a and 14b) and total number of infected individuals infected with HPV and persistent HPV, respectively (Figures 14c and 14d), when the universal control strategy is implemented and when there is no control administered. Here, *β*_s_ = 7.0, *β*_H_ = 2.0. All other parameters as in Table 2

**Figure 15:**
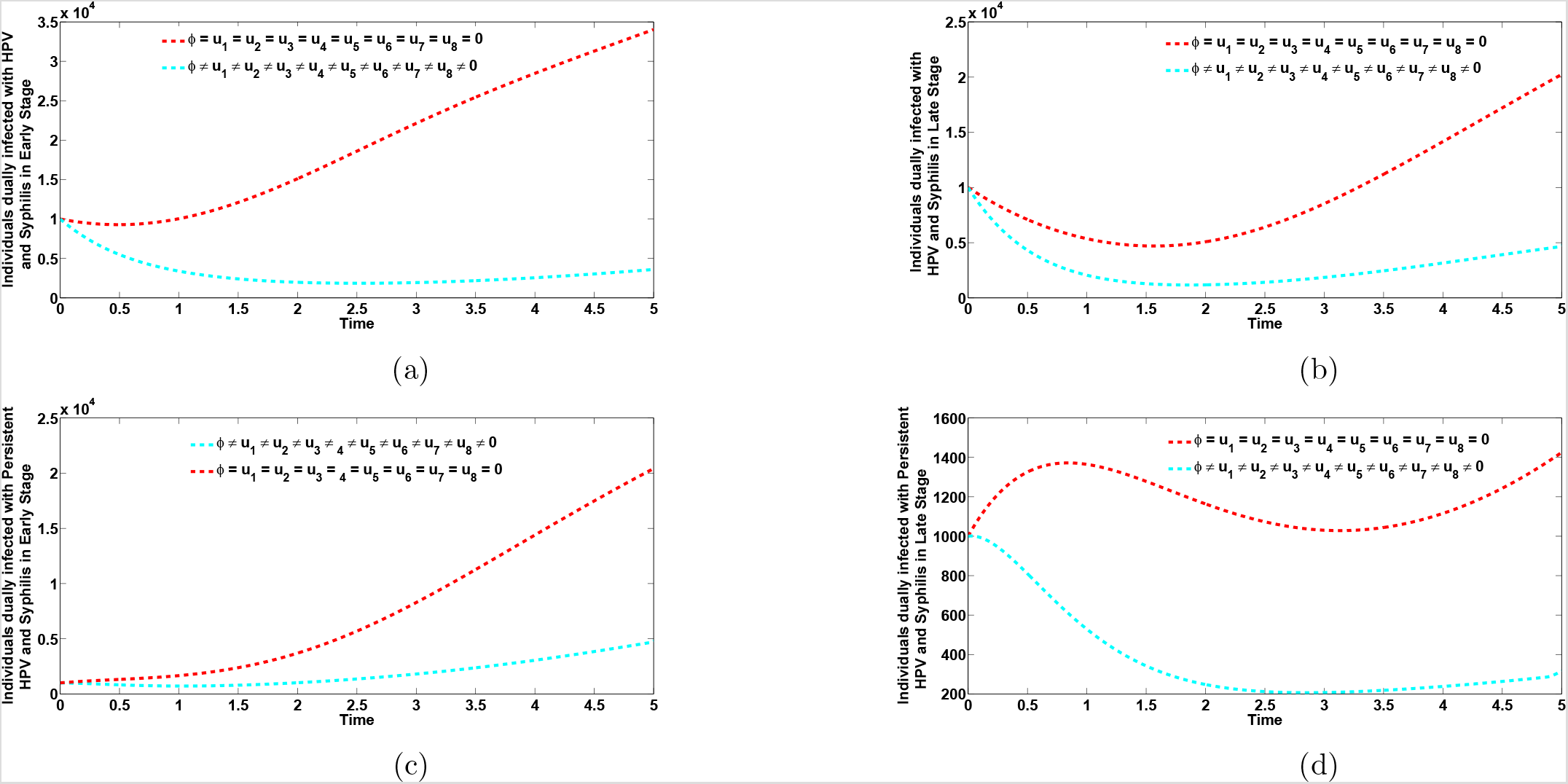
Plots of the co-infection cases for individuals dually infected with HPV and syphilis in early and late stages of infection, respectively (Figures 15a and 15b) and co-infection cases for individuals dually infected with persistent HPV and syphilis in early and late stages of infection, respectively (Figures 15c and 15d), when the universal control strategy is implemented and when there is no control administered. Here, *β*_s_ = 7.0, *β*_H_ = 2.0. All other parameters as in Table 2

**Figure 16:**
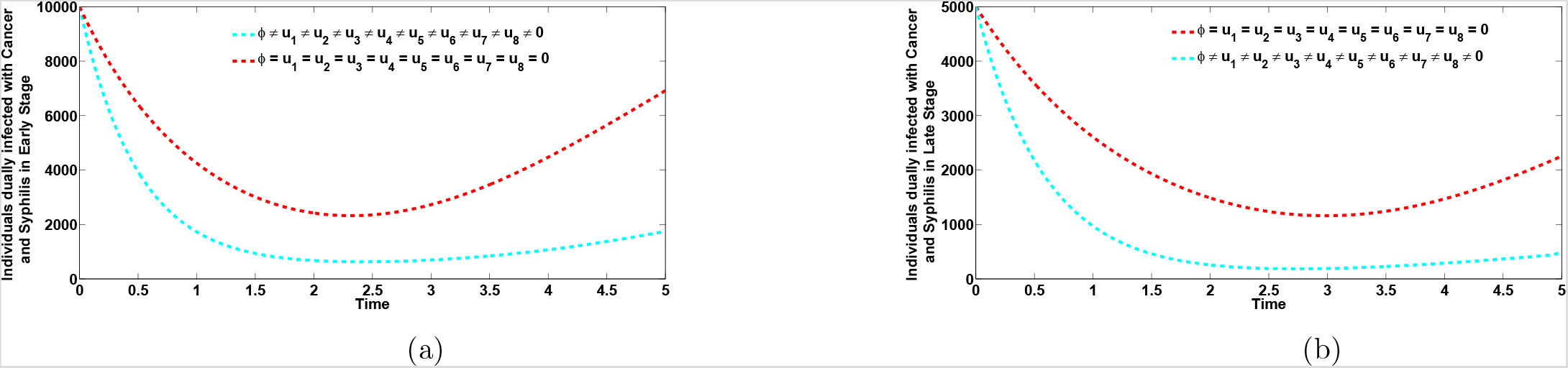
Plots of the co-infection cases for individuals dually infected anal cancer and syphilis in early and late stages of infection, respectively (Figures 16a and 16(b), when there is universal control strategy and when there is no control administered. Here, *β*_s_ = 7.0, *β*_H_ = 2.0. All other parameters as in Table 2

### 5.2 Cost-effectiveness analysis

The cost-effectiveness analysis is used to evaluate the health interventions related benefits so as to justify the costs of the strategies [5]. This is obtained by comparing the differences among the health outcomes and costs of those interventions; achieved by computing the incremental cost-effectiveness ratio (ICER), which is defined as the cost per health outcome. It is given by:

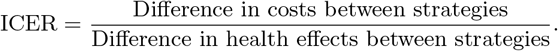

We calculated the total number of co-infection cases averted and the total cost of the strategies applied in Table 3. This is equally presented inFigure 18. The total number of co-infection cases prevented is obtained by calculating the total number of individuals when controls are implemented and the total number when there is no control applied. Similarly, we apply the cost functions 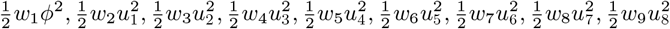 over time, to compute the total cost for the various strategies implemented. We compared the cost-effectiveness of strategy I (Optimal HPV vaccination strategy for sexually active susceptible individuals) and strategy II (syphilis treatment controls for singly infected individuals).

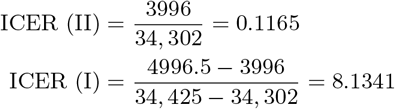

**Table 3:** Increasing order of the total infection averted due to the control strategies.

| Strategy | Total infection averted | Total cost | ACER | ICER |
| --- | --- | --- | --- | --- |
| II: $u_1(t), u_2(t)$ | 34, 302 | 3, 996 | 0.1165 | 0.1165 |
| I: $\phi(t)$ | 34, 425 | 4, 996.5 | 0.1451 | 8.1341 |
| III: $u_3(t), u_4(t), u_5(t), u_6(t), u_7(t), u_8(t)$ | 41, 372.6 | 5, 954.5 | 0.1439 | 0.1379 |
| IV: $\phi(t), u_1(t), u_2(t), u_3(t), u_4(t), u_5(t), u_6(t), u_7(t), u_8(t)$ | 69, 380 | 10, 286 | 0.1483 | 0.1547 |

**Figure 17 Figure 18:**
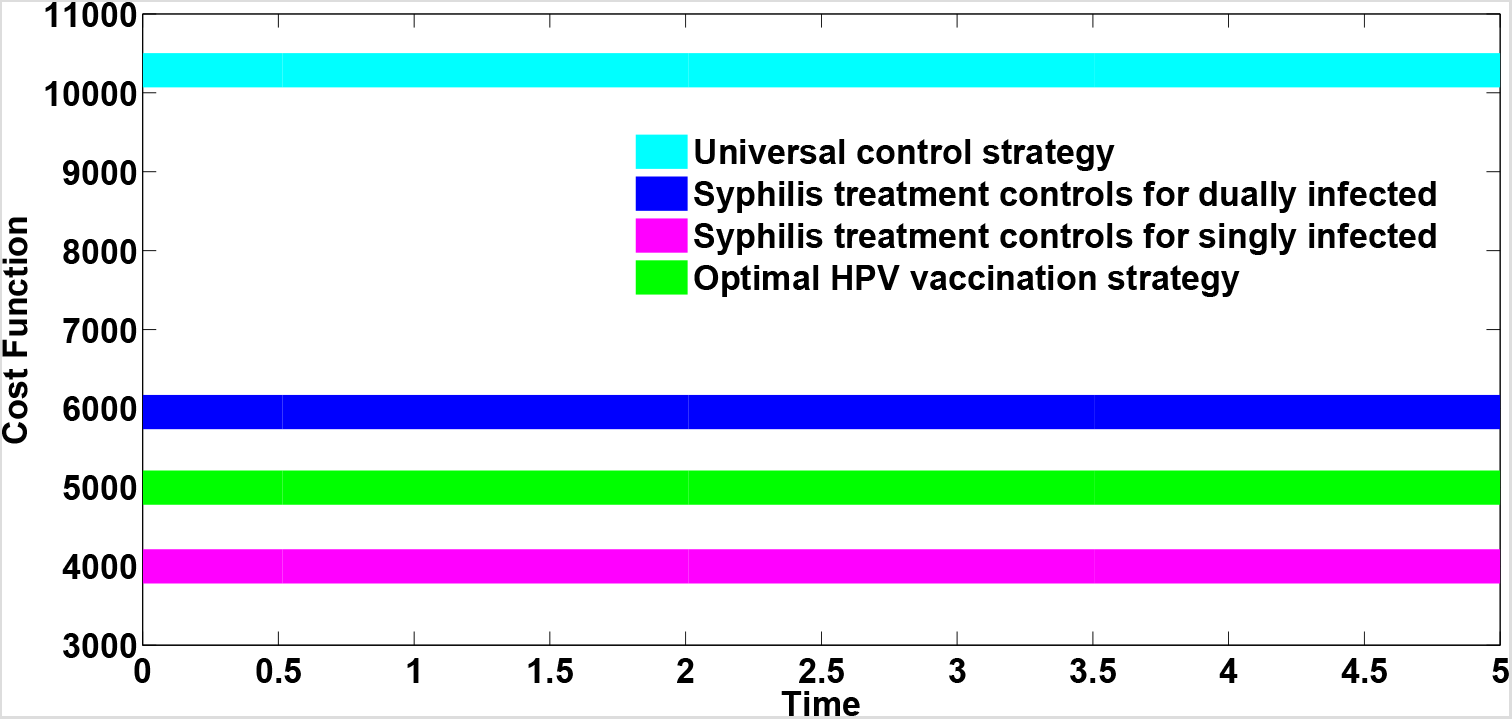
Cost functions of the different control strategies

From ICER (I) and ICER(II), we see a cost saving of 0.1165 observed for strategy II over strategy I. This implies that strategy I strongly dominated strategy II, showing that strategy I is more costly and less effcetive compared to strategy II. Hence, strategy I is removed from subsequent ICER computations, shown in Table 4. We now compare strategy II and strategy III.

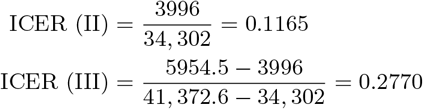

**Table 4:**
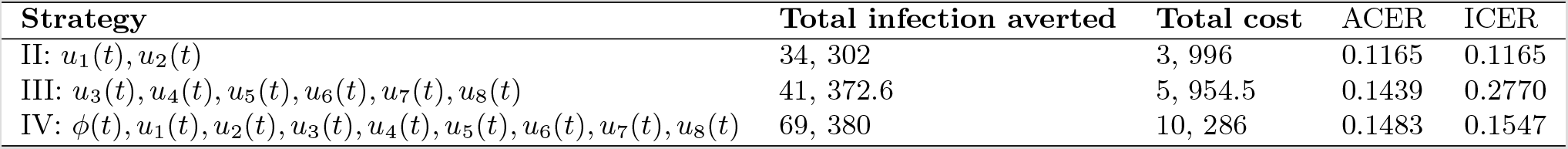
Increasing order of the total infection averted due to the control strategies.

Comparing strategy II and strategy III, we observe that ICER (III) is greater than ICER (II), showing that strategy III strongly dominated strategy II and is more expensive and less effective compared to strategy II. Therefore, strategy III is removed from the list of next alternative strategies and we re-calculate ICER for the remaining competing strategies II and IV, as shown in Table 5.

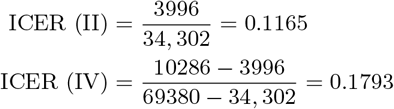

**Table 5:**
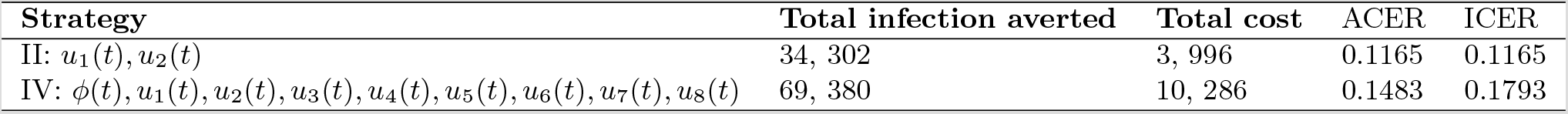
Increasing order of the total infection averted due to the control strategies.

| Strategy | Total infection averted | Total cost | ACER | ICER |
| --- | --- | --- | --- | --- |
| II: $u_1(t), u_2(t)$ | 34, 302 | 3, 996 | 0.1165 | 0.1165 |
| IV: $\phi(t), u_1(t), u_2(t), u_3(t), u_4(t), u_5(t), u_6(t), u_7(t), u_8(t)$ | 69, 380 | 10, 286 | 0.1483 | 0.1793 |

Comparing strategy II and strategy IV, it can be observed that ICER (IV) is greater than and strongly dominates ICER (II), showing that strategy IV is more costly and less effective compared to strategy III. As a result, strategy II (the strategy that implements syphilis treatment controls for singly infected individuals) has the least ICER and is the most cost-effective of all the control strategies for the control of HPV and syphilis co-infections. This is clearly illustrated in Figure 18, which also agrees with the results obtained from both ACER and ICER methods that strategy II is the most cost-effective strategy.

## 6 Conclusion

In this work, we have developed and presented a co-infection model for HPV and syphilis with cost-effectiveness optimal control analysis. The full co-infection model was shown to undergo the phenomenon of backward bifurcation when a certain condition was satisfied. The global asymptotic stability of the disease-free equilibrium of the full model was shown not to exist, when the associated reproduction number was less than unity. The existence of endemic equilibrium of the syphilis-only sub-model was shown to exist and the global asymptotic stability of the disease-free and endemic equilibria of both the syphilis-only sub-model and HPV-only sub-model were established. The global asymptotic stability of disease-free equilirbium of the HPV-only sub-model was also proven.

Numerical simulations of the optimal control model showed that:

i. HPV vaccination control has a positive population level impact in reducing the burden of HPV and the co-infection cases in a population
ii. Syphilis treatment controls for singly infected individuals not only help bring down the burden of syphilis infection, but also reduce the burden of the HPV and syphilis co-infections.
iii. The control strategy which implements syphilis treatment for singly infected individuals is the most cost-effective of all the control strategies in reducing the burden of HPV and syphilis co-infections.

## Data Availability

Not applicable

http://www.cdc.gov/vaccines/programs/vfc/awardees/vaccine-management/price-list

## References

[1] Aadland D, Finnoff D, Huang K, syphilis cycles. J Econ Financ (2013) 13(1): 297–348

[2] Adams RA, Calculus: A Complete Course, Pearson Addison Wesley, Toronto, 2006.

[3] Ahmed HG, Bensumaidea SH, Ashankyty IM, Frequency of human papillomavirus (HPV) subtypes 31, 33, 35, 39 and 45 among Yemeni women with cervical cancer, Infect. Ag. Cancer 10 (29) (2015) 1–6.

[4] Alsaleh AA, Gumel AB, Analysis of a risk-structured vaccination model for the dynamics of oncogenic and warts-causing HPV types, Bull. Math. Biol. Vol. 76 (2014) 1670–1726.

[5] Agusto FB, Adekunle AI, Optimal control of a two-strain tuberculosis-HIV/AIDS co-infection model, BioSystems, (2014) (119) 20–24.

[6] Andrawus J, Nwankwo A, Okuonghae D, Bifurcation analysis of a mathematical model for TB-Dengue co-infection. Nig. Research J. Engineering and Environmental Sciences, (2017)2 (2): 390–407.

[7] Andrawus J, Nwankwo A, Okuonghae D, Population dynamics of a mathematical model for TB-Dengue co-infection. Transactions of the Nig. Ass. of Math. Physics, (2017) 5: 285–292.

[8] Berman SM, Maternal syphilis: pathophysiology and treatment, Bull. World Health Organ. 82 (6) (2004) 433–438.

[9] Brazil Demographics Profile, 2018, Available at http://www.indexmundi.com/brazil/demographics_profile. Accessed: 31 December, 2018.

[10] Castillo-Chavez C, Song B, Dynamical models of tuberculosis and their applications, Mathematical Biosci. Engnrg. Vol. 2 (2004) 361–404.

[11] Castillo-Chavez C, Feng Z, Huang W, On the computation of *R*_0_ and its role on global stability, in Mathematical Approaches for Emerging and Reemerging Infectious Diseases: An Introduction (Minneapolis, MN, 1999), 229-250, IMA Vol. Math. Appl., 125 Springer New York.

[12] CDC,2015.CDC Vaccine Price List, Vaccines for Children Program (VFC). http://www.cdc.gov/vaccines/programs/vfc/awardees/vaccine-management/price-list/.

[13] Daling JR, Weiss NS, Klopfenstein LL, Cochran LE, Chow WH, Daifuku R, Correlates of homosexual behaviour and the incidence of anal cancer. JAMA (1982) 247: 1988–1990.

[14] Iboi E, Okuonghae D, Population dynamics of a mathematical model for syphilis, Applied Mathematical Modelling, Vol. 40 (2016) 3573–3590.

[15] Ghosh M, Olaniyi S, Obabiyi OS, Mathematical analysis of reinfection and relapse in malaria dynamics, Applied Mathematics and Computation, (2020) 373 1–18.

[16] Garnett GP, Aral SO, Hoyle DV, Cates W, Anderson RM, The natural history of syphilis: implications for the transition dynamics and control of infection. Sex Transm Dis (1997) 24(4): 185–200

[17] Grassly C, Fraser C, Garnett GP, Host immunity and synchronized epidemics of syphilis across the United States. Nature (2005) 433: 417–421

[18] Kahn JG, Jiwani A, Gomez GB, Hawkes SJ, Chesson HW, (2014) The Cost and Cost-Effectiveness of Scaling up Screening and Treatment of Syphilis in Pregnancy: A Model. PLoS ONE 9(1): e87510. doi: 10.1371/journal.pone.0087510.

[19] La Salle J, Lefschetz S, The Stability of Dynamical Systems, SIAM, Philadelphia, 1976.

[20] Lenhart S, Workman JT, 2007. Optimal Control Applied to Biological Models. Chapman & Hall, Boca Raton.

[21] Malik MT, Reimer J, Gumel AB, Elbasha EH, Mahmud SM, The impact of an imperfect vaccine and pap cytology screening on the transmission of Human Papillomavirus and occurrence of associated cervical dysplasia and cancer, Math. Biosci. Engrg., Vol. 10(4) (2013).

[22] Malik T, Imran M, Jayaraman R, Optimal control with multiple human papillomavirus vaccines, Journal of Theoretical Biology, (393) 179–193, 2016.

[23] Milner F, Zhao R, A new mathematical model of syphilis. Math Model Nat Phenom (2010) 5(6): 96–108.

[24] Nwankwo A, Okuonghae D, Mathematical analysis of the transmission dynamics of HIV syphilis Co-infection in the presence of treatment for syphilis, Bull. Math. Biol. 80 (3) (2018) 437–492.

[25] Miranda AE, Figueiredo NC, Pinto VM, Kimberly P, Talhari S, Risk factors for syphilis in young women attending a family health program in Vitória, Brazil, An. Bras. Dermatol. 2012; 87(1): 76–83.

[26] da Motta LR, Sperhacke RD, Adami Ad-G, Pharma B, Kato SK, Vanni AC, Paganella MP, de Oliveira MCP, Giozza SP, da Cunha ARC, Pereira GFM, Benzaken AS, Medicine (2018) 97: 47

[27] Okuonghae D, Gumel AB, Ikhimwin BO, Iboi E, Mathematical Assessment of the Role of Early Latent Infections and Targeted Control Strategies on Syphilis Transmission Dynamics. Acta Biotheor (2019) 67 47–84. https://doi.org/10.1007/s10441-018-9336-9

[28] Okuonghae D, Omame A (2020) Analysis of a mathematical model for COVID-19 population dynamics in Lagos, Nigeria, Chaos Solitons Fractals 139: 110032

[29] Omame A, Umana RA, Okuonghae D, Inyama SC, Mathematical analysis of a two-sex Human Papillomavirus (HPV) model, International Journal of Biomathematics, 11 (2018)(7)

[30] Omame A, Okuonghae D, Umana RA, Inyama SC, Analysis of a co-infection model for HPV-TB, Applied Mathematical Modelling, 77 (2020) 881–901.

[31] Omame A, Okuonghae D, Inyama SC, A mathematical study of a model for HPV with two high risk strains, in Mathematics Applied to Engineering, Modelling, and Social Issues Studies in Systems, Decision and Control (2020) 200, F. Smith, H. Dutta and J. N. Mordeson (eds.)

[32] Omame A, Sene N, Nometa I, Nwakanma CI, Nwafor EU, Iheonu NO, Okuonghae D, Analysis of COVID-19 and comorbidity co-infection Model with Optimal Control, medRxiv preprint doi: https://doi.org/10.1101/2020.08.04.20168013.

[33] L.S. Pontryagin, V.G. Boltyanskii, R.V. Gamkrelidze, E.F. Mishchenko, The Mathematical Theory of Optimal Processes, Wiley, New York, 1962

[34] Souza LMS, Miller WM, Nery JAC, de Andrade AFB, Asensi MD, A syphilis Co-Infection Study in Human Papilloma Virus Patients Attended in the Sexually Transmitted Infection Ambulatory Clinic, Santa Casa de Misericórdia Hospital, Rio de Janeiro, Brazil, The Brazilian Journal of Infectious Diseases 2009 13(3): 207–209

[35] Tseng H-F, Morgenstern H, Mack TM, Peters RK, Risk factors for anal cancer: results of a population-based case:control study, Cancer Causes and Control (2003) 14: 837–846

[36] Saad-Roy CM, Shuai Z, van den Driessche P, A mathematical model of syphilis transmission in an MSM population. Math Biosci. (2016) 277: 59–70

[37] Saldana F, Korobeinikov A, Barradas I, Optimal Control against the Human Papillomavirus: Protection versus Eradication of the Infection, Abstract and Applied Analysis, https://doi.org/10.1155/2019/4567825

[38] Saslow D, Solomon D, Lawson HW, American Cancer Society, American Society for Colspcopy and Cervical Pathology, and American Society for Clinical pathology screening guidelines for the prevention and early detection of cervical cancer, American Journal of Clinical Pathology, (2012) 137 516–542.

[39] Soares CC, George I, Lampe E, Lewis L, Morgado MG, HIV-1, HBV, HCV, HTLV, HPV-16/18, and Treponema pallidum Infections in a Sample of Brazilian Men Who Have Sex with Men. PLoS ONE (2014) 9(8): e102676. doi: 10.1371/journal.pone.0102676

[40] Umana RA, Omame A, Inyama SC (2016) Deterministic and Stochastic Models of the Dynamics of Drug Resistant Tuberculosis; FUTO Journals Series, 2(2): 173–194.

[41] Uwakwe JI, Inyama SC, Omame A (2020) Mathematical Model and Optimal Control of New-Castle Disease (ND). Applied and Computational Mathematics. 9(3): 70–84. doi: 10.11648/j.acm.20200903.14

[42] P. van den Driessche and J. Watmough, Reproduction numbers and sub-threshold endemic equilibria for compartmental models of disease transmission, Math. Biosci, Vol. 180 (2002), 29–48.

[43] van Damme P, Bonanni P, Bosch X, Joura E, Kjaer SK, Meijer CJLM, Petry K-U, Soubeyrand B, Verstraeten T, Stanley M, Use of the nonavalent HPV vaccine in individuals previously fully or partially vaccinated with bivalent or quadrivalent HPV vaccines, Vaccine, 34 (2016) 757–761.

[44] World Health Organization (2019) Eliminating congenital syphilis. http://www.who.int/reproductive-health/stis/syphilis.html. Accessed 6 August 2019.

[45] Zhang X, Yu J, Li M, Sun X, Han Q, Li M, Zhou F, Li X, Yang Y, Xiao D, Ruan Y, Jin Q, Gao L, Prevalence and Related Risk Behaviors of HIV, syphilis, and Anal HPV Infection Among Men who have Sex with Men from Beijing, China, AIDS Behav (2013) 17: 1129–1136

